# The risk of severe COVID-19 and mortality from COVID-19 in people living with HIV compared to individuals without HIV - a systematic review and meta-analysis of 1 268 676 individuals

**DOI:** 10.1101/2021.07.03.21259958

**Authors:** Lovemore Mapahla, Asmaa Abdelmaksoud, Rida Arif, Nazmul Islam, Albert Chinhenzva, Suhail A. R. Doi, Tawanda Chivese

**Affiliations:** Division of Epidemiology and Biostatistics, Department of Global Health, Faculty of Medicine and Health Sciences, Stellenbosch University, Cape Town, South Africa; Department of Population Medicine, College of medicine, QU Health, Qatar University, Doha, Qatar; Department of Public Health, College of Health Sciences, QU Health, Qatar University, Doha, Qatar

**Author notes:** Corresponding author: Tawanda Chivese, Department of population medicine, college of medicine, QU Health, Qatar University, Doha, Qatar.

**Keywords:** HIV, COVID19, mortality, hospitalization, intensive care services

## Abstract

**Background:** There is conflicting evidence about the risk of mortality and severe disease due to COVID-19 in people living with HIV (PLHIV).

**Objectives:** To compare mortality, hospitalization, and the need for intensive care services due to COVID-19 between PLHIV and individuals without HIV based on data from the existing literature.

**Methods:** A comprehensive search in PubMed, Cochrane Library, Scopus, China Academic Journals Full Text Database, the Database of Abstracts of Reviews of Effectiveness (DARE) and and the medRXIV and bioRxiv databases of preprints was carried out. Each data source was searched from 1 January 2020 to 20^th^ of February 2021. Eligible studies were case control, cross-sectional and cohort studies where participants had confirmed COVID-19. From each study, data on numbers of PLHIV and individuals without HIV for each outcome were extracted. Study quality was assessed using the MethodologicAl STandard for Epidemiological Research (MASTER) scale. Data synthesis used a bias adjusted model and predefined age and geographical subgroups were analysed.

**Results:** Of a total of 2757 records identified, 11 studies, from 4 countries, the United Kingdom, Spain, the United States of America and South Africa, were included. The total participants assessed for the outcomes in this meta-analysis were 1 268 676 of which 13 886 were PLHIV. Overall, the estimated effect of HIV on mortality suggested some worsening (OR 1.3, 95% CI: 0.9 – 2.0, I^2^ = 78.6%) with very weak evidence against the model hypothesis at this sample size. However, in individuals aged <60 years, the estimated effect on mortality suggested more worsening in PLHIV (OR 2.7, 95% CI: 1.1 - 6.5, I^2^ = 95.7%) with strong evidence against the model hypothesis at this sample size. HIV was also associated with an estimated effect on hospitalization for COVID-19 that suggested worsening (OR 1.6, 95% CI: 1.3-2.1, I^2^ = 96.0%) also with strong evidence against the model hypothesis at this sample size. A secondary analysis of the included studies suggested no difference, by HIV status, in the prevalence of pre-existing conditions.

**Conclusion:** People living with HIV have higher risk of death and hospitalisation from COVID-19, compared to individuals without HIV. A secondary analysis suggests this is not due to associated comorbid conditions. The difference in mortality is exaggerated in those younger than 60 years of age.

**Registration:** PROSPERO: CRD42020221311 (https://www.crd.york.ac.uk/prospero/display_record.php?RecordID=221311)

**Evidence before this study:** Findings from existing studies have shown conflicting evidence concerning the risk of severe COVID-19 and death from COVID-19 in people living with HIV (PLHIV) compared to people without HIV. Evidence from three existing systematic reviews suggests that the risk of severe COVID-19 and death from COVID-19 in PLHIV may be similar to that in individuals without HIV. However, findings from three large cohort studies and one meta-analysis of four studies suggest that the risk of death from COVID-19 in PLHIV may be higher than that in individuals without HIV. One of the large cohort studies, which is also included in the previous meta-analysis, consisted of individuals with unknown COVID-19 status, and therefore there is still debate concerning the risk of severe COVID-19 outcomes in PLHIV.

**Added value of this study:** In this meta-analysis of 11 studies with 1 268 676 individuals with confirmed COVID-19, we found a stronger difference in mortality by HIV status for those individuals below the age of 60 years, and over this age, HIV had an attenuated effect on mortality, suggesting that age-related mortality overshadows PLHIV related mortality. Further, PLHIV had increased odds of being hospitalized and needing intensive cares services, probably related to increased COVID-19 severity in PLHIV. A secondary analysis of the included studies suggested no difference in the prevalence of pre-existing conditions.

**Implications of all the available evidence:** Our findings suggest that PLHIV are at higher risk than the general population and should be prioritized for vaccine coverage and monitoring if diagnosed with COVID-19. This is especially important for countries in Sub-Saharan Africa that have a high burden of HIV in the younger populations who are more vulnerable.

**Strengths:** This study was carried out rigorously following the PRISMA guidelines for systematic reviews and meta-analyses. We used a comprehensive search strategy across most of the main citation databases to ensure that no relevant studies were missed. We included studies where participants had confirmed COVID-19 only and we synthesized the findings from studies using a bias adjustment model that took into consideration the quality of included studies.

**Limitations:** All studies included in this review are observational studies and conclusions about causality require cautious interpretation. Due to a lack of data from included studies, we were not able to analyse the effect of being on treatment for HIV, and HIV control variables such as viral load and CD4 counts on COVID-19 hospitalization, intensive care services and mortality. Lastly most of the included studies had small samples overall or for PLHIV and this may affect the effect estimates in this analysis. Future research is therefore indicated to confirm these findings.

## Introduction

The number of individuals with confirmed COVID-19 continues to increase globally (1). Identifying groups of people who are susceptible to severe COVID-19 and death from COVID-19 is a priority, as these groups may need additional protections and prioritization for vaccination against COVID-19 (2, 3). There are indications that people living with HIV (PLHIV) may be one such vulnerable group due to persistent immune-suppression, although data from published studies remain inconclusive (4). As of 2020 at least 38 million people were living with HIV globally, more than two-thirds of PLHIV being in Sub-Saharan Africa (5). Since December 2020, the number of confirmed cases of COVID-19 has increased in Sub-Saharan Africa (1), where health systems are ill equipped to treat surges in the need for hospitalization, mechanical ventilation and intensive care services associated with severe COVID-19. The COVID-19 pandemic has also affected several aspects of the HIV care cascade and may erode some of the gains made in the fight against HIV (6). It is imperative to quantify the risk that COVID-19 poses to PLHIV, as this may help preparedness planning for countries that are disproportionately affected by HIV.

Evidence from several systematic reviews (4,7–10) and individual studies (11–14) suggested that PLHIV does not pose an additional risk of mortality from COVID-19. In fact, it has been hypothesized that most PLHIV who are adherent to antiretroviral therapy (ART) may have some degree of protection against severe COVID-19 because some ART such as nucleoside reverse transcriptase inhibitors, like tenofovir disoproxil fumarate (TDF) and lamivudine, have shown some degree of efficacy against the SARS-CoV-2 virus*, in-vitro*, by inhibiting RNA-dependent RNA polymerase (RNAdRNAp) (14, 15). However, this has not been proven by findings from clinical research.

Emerging evidence (16–19) suggests that individuals living with HIV may have a higher risk for severe COVID-19 compared to individuals without HIV, partly attributed to a higher prevalence of comorbidities in PLHIV which are associated with severe COVID-19, such as diabetes and hypertension. Several studies (16–18) have been published and their findings suggest that PLHIV may have a higher risk of severe COVID-19 and mortality from COVID-19, compared to individuals without HIV, although estimates of the risk are different. Bhaskaran and colleagues (18) analysed data from 17.3 million individuals in the primary care database, OPENSAFELY, and reported an adjusted hazard ratio of 2.6 (95%CI 1.7 – 3.8) for mortality in PLHIV compared to individuals without HIV in the United Kingdom. A major limitation of this study is that they used population totals to infer risk as they did not have the totals with confirmed COVID-19. In the USA, a population-based study of roughly 20 million individuals (17) reported a modest adjusted standardized rate ratio of 1.2 (95%CI 1.1 – 1.4) for mortality due to COVID-19 in PLHIV compared to individuals without COVID-19. Findings from this study also suggested that PLHIV are more likely to be hospitalized for COVID-19, compared to individuals without HIV, implying a more severe disease course in PLHIV. Another population-based study (16), this time from South Africa, analysed data of 3.5 million public sector patients and reported an adjusted hazard ratio of 2.1 (95%CI 1.7 – 2.7) for mortality in PLHIV compared to individuals without HIV. A recent meta-analysis (19) reported a pooled risk ratio for mortality from COVID-19 of 1.8 (95%CI 1.2 – 2.6) although they included only four studies for this outcome and used an effect measure that probably should not be used in meta-analysis (21). Many of the existing reviews have also not investigated the risk of other key clinical severity outcomes for COVID-19, hospitalisation and the need for intensive care services in PLHIV. To date, more than ten studies have been published with data comparing severity and mortality from COVID-19 between PLHIV and individuals without HIV. A meta-analysis of these studies may help update the conclusions reached thus far for the risk of COVID-19 related outcomes in PLHIV. This study therefore updates previous meta-analyses and used more robust methods to define the risk status in PLHIV. In secondary analyses, we compared the prevalence of comorbid conditions in individuals with COVID-19 by HIV status.

## Methods

### Study Design and protocol registration

The design and conduct of this study followed the Preferred Reporting Items for Systematic Reviews and Meta-Analysis (PRISMA) guidelines (20) and the protocol was registered with the international database of prospectively registered protocols of systematic reviews and meta-analyses (CRD42020221311).

### Search strategy and data sources

We searched for studies in the following electronic databases; PubMed, Cochrane Library, Scopus, China Academic Journals Full Text Database, the Database of Abstracts of Reviews of Effectiveness (DARE) and and the medRXIV and bioRxiv databases of preprints. We manually screened all references of all included studies for any additional studies which could have been missed. We also carried out a citation search of the top 20 similar articles of all included studies on PubMed to retrieve studies that might have been missed in the original electronic search. Each data source was searched between 1 Jan 2020 and 20^th^ of February 2021. The following search terms for COVID-19 were used: “COVID19” OR “Coronavirus” OR “novel coronavirus” OR “SARS-CoV-2” OR “COVID” OR “COVID-19”. We used the following search terms for HIV “HIV” OR “Hiv” OR “hiv” OR “human immunodeficiency virus” OR “arv” OR “ARV” OR “controlled HIV” OR “uncontrolled HIV” OR PLHIV OR ART. The full electronic search strategy is given in supplementary Table 1.

**Table 1.**
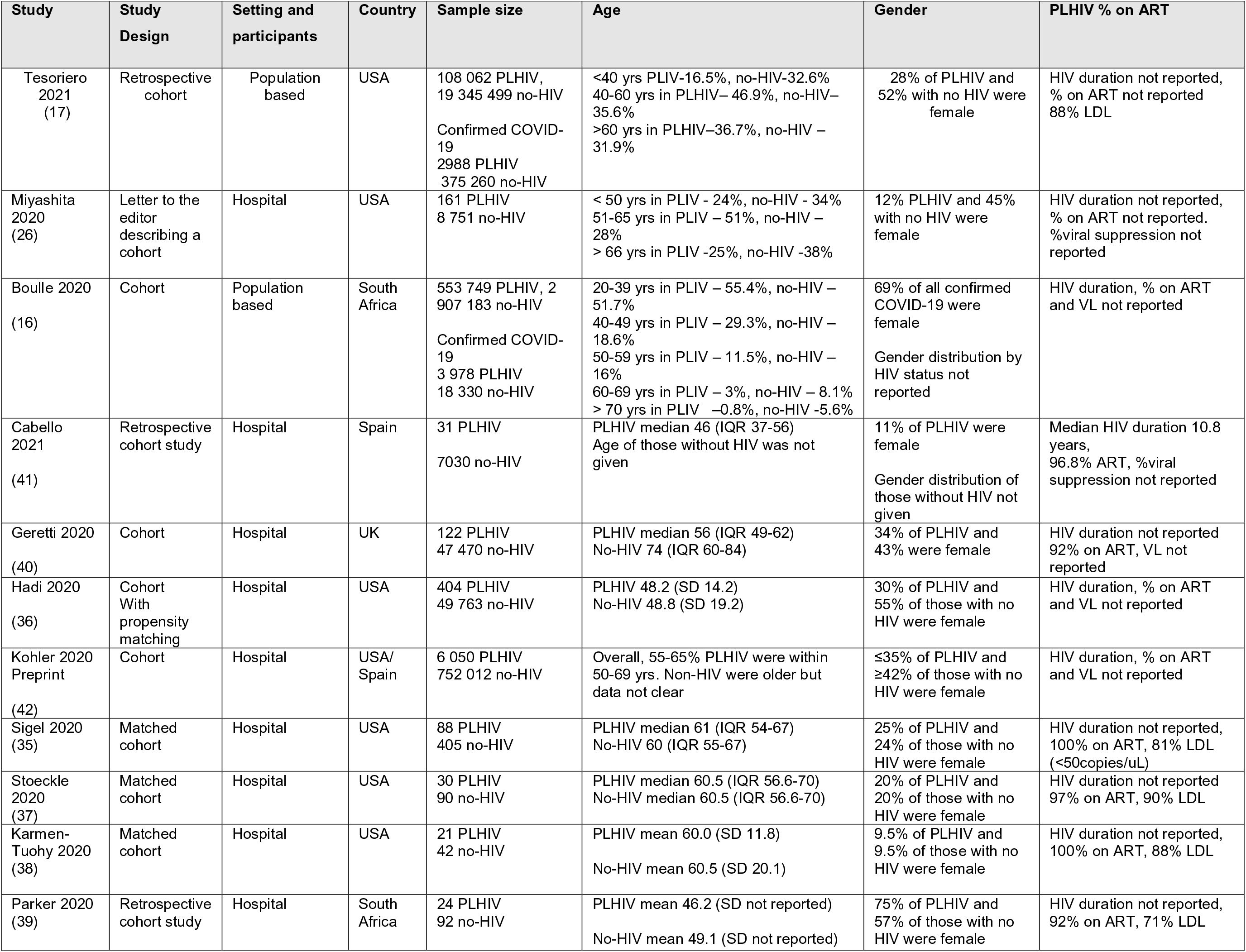
Characteristics of included studies

### Screening of studies for inclusion

Study records identified from the searches were exported into Endnote referencing software for removal of duplicates and then exported into the Rayyan platform (22) for selection. Six reviewers blindly screened titles and abstracts of studies uploaded on Rayyan platform Rayyan (http://rayyan.qcri.org/) (22) and conflicts were resolved through discussion. The studies included after screening of abstracts and titles were then assessed for eligibility using full text.

### Eligibility

The full text of the studies from Rayan were assessed by two reviewers per study using predefined inclusion and exclusion criteria. Observational studies including case-control, cohort and cross-sectional studies were eligible for inclusion in this systematic review if they compared mortality, hospitalization or the need for intensive care services due to COVID-19 between PLHIV and people without HIV. Case studies, case series, duplicate reports, commentaries and reviews were excluded. Studies which did not compare the outcomes of interest between PLHIV and individuals without HIV were excluded. Studies which did not report on the outcomes of interest of this review were also excluded.

### Outcomes

The primary outcome of interest was mortality in individuals with COVID-19, either measured as all-cause mortality, 28-day mortality, 30-day mortality or COVID-19 specific mortality. Secondary outcomes were hospitalization and the need for intensive care services for COVID-19. The need for intensive care services was defined as any of; transfer to the intensive care unit (ICU) and/or intubation and/or mechanical ventilation. We also extracted data on pre-existing diseases by HIV status, in the included studies where data were available. The comorbidities of interest were diabetes, hypertension, cardiovascular diseases, obesity (body mass index≥ 30kg/m2), dyslipidaemia, chronic kidney disease, malignancies, and asthma.

### Data extraction

Two reviewers independently extracted data from each included study. The extracted information included study characteristics such as author, year of publication, study design, study outcome; study population: country of study, study setting, sample size; and the number of participants with COVID-19, if the number differed from the sample size. We also extracted the following outcomes, by HIV status; the number of participants hospitalised for COVID-19, the number of participants transferred to the intensive care unit (ICU) and/or requiring intubation and/or mechanical ventilation and the number of deaths. We combined data for ICU admission, intubation and mechanical ventilation, and grouped these as intensive care services.

Where studies reported population-based numbers of PLHIV and individuals without HIV for risk estimates, we extracted data for individuals with confirmed COVID-19 only.

To compare the prevalence of chronic comorbidities between PLHIV with COVID-19 and individuals without HIV with COVID-19, we extracted data on the number of individuals with each comorbidity in the two groups. Studies that used matched designs were excluded from this analysis unless unmatched data were available.

### Assessment of study quality

Two authors independently assessed the methodological quality of studies, for the main outcome, mortality, across safeguards listed within 7 standards using the MethodologicAl STandard for Epidemiological Research (MASTER) scale (23). Any disagreements between two reviewers were solved through discussion. The MASTER scale consists of seven safeguards standards, namely, equal recruitment (items 1-4), equal retention (items 5-9), equal ascertainment (items 10-16), equal implementation (items 17-22), equal prognosis (items 23-28), sufficient analysis (items 29-31) and temporal precedence (items 32-36). The tool is more comprehensive than and includes all the items in the Cochrane risk of bias tool. Each study gets a safeguard count out of 36, higher counts indicating higher study quality. In this review, this count was then used in a quality-adjusted meta-analysis after conversion to quality ranks.

### Synthesis of findings

The characteristics of the included studies are shown in Table 1 and described narratively. Tableau software (24) was used to create a map showing the geographic origin of the included studies. We also used tables to summarize findings from studies on each of the outcomes of hospitalization, intensive care services and mortality. For each study we reported the study’s adjusted odds ratios in the tables and then summarized the findings from all the studies per outcome in the text.

In pooled analyses, we recalculated unadjusted odds ratios for risk of each outcome of hospitalization, intensive care services and mortality in PLHIV compared to individuals without HIV, from each study, because the studies reported different effect measures across all the outcomes. We used the quality effects model to pool the results of studies for each outcome (25). Th assumption of this quality effects model is that there is one true effect size shared by all included studies and any deviations from this due to random and/or systematic error. Results of the meta-analysis were presented in forest plots and pooled odds ratios (OR) and their 95% confidence intervals (95%) was the effect size reported. We used forest plots without the pooled estimates to present the results from the studies.

We pre-specified subgroup analysis for the region of origin of the studies (sub-Saharan Africa, Europe and North America) and for age groups, using the broad categories of age<60 years and ≥60 years. In the age-group analysis, we also included a study (26) where the authors reported data as <66 years and ≥66 years. To compare the prevalence of chronic comorbidities between PLHIV and individuals without HIV, we also used the quality effects models with weights from the MASTER quality assessment.

The results of a sensitivity analysis by the random effects model were presented in the supplementary material.

Heterogeneity was assessed using the Cochran’s Q test (27) and quantified heterogeneity using the I^2^ statistic which is the between-study variance as a percentage of total variance (27). Heterogeneity ranges from no heterogeneity when I^2^ is 0% to high heterogeneity when I^2^ is more than 75%. Values above 50%, but less than or equal to 75% are described as substantial heterogeneity (27).

Doi plots and the LFK index were used for assessing publication bias as they are more easier to interpret (28) than funnel plots and in this case funnel plots could not be used as they are not recommended when there are less than 10 studies per outcome in a meta-analysis (29). An LFK index with a magnitude greater than one reflects increasing asymmetry and a higher likelihood of publication bias (38). Stata version 15.1 (30) *metan* module was used for the meta-analysis.

### Ethics

This systematic review and meta-analysis used aggregate data from already published studies therefore no ethical approval was required.

## Results

### Search results

A total of 2757 records were identified from searches of both electronic databases and additional citation searches, and 1746 records were assessed for inclusion, using the title and abstract only, after removal of duplicates (Fig. 1). From these 1746 studies, 32 studies were selected for full text screening, and 21 studies were subsequently excluded with reasons after full text screening. Most of the 21 studies were excluded because they had no non-HIV comparison group (n= 16), three were excluded because they did not report on the outcomes of this review (31–33), and one study was a duplicate (34). One study (19) used population-based totals and did not report totals with confirmed COVID-19 infection and was therefore excluded. A total of 11 studies were finally included.

**Fig.1:**
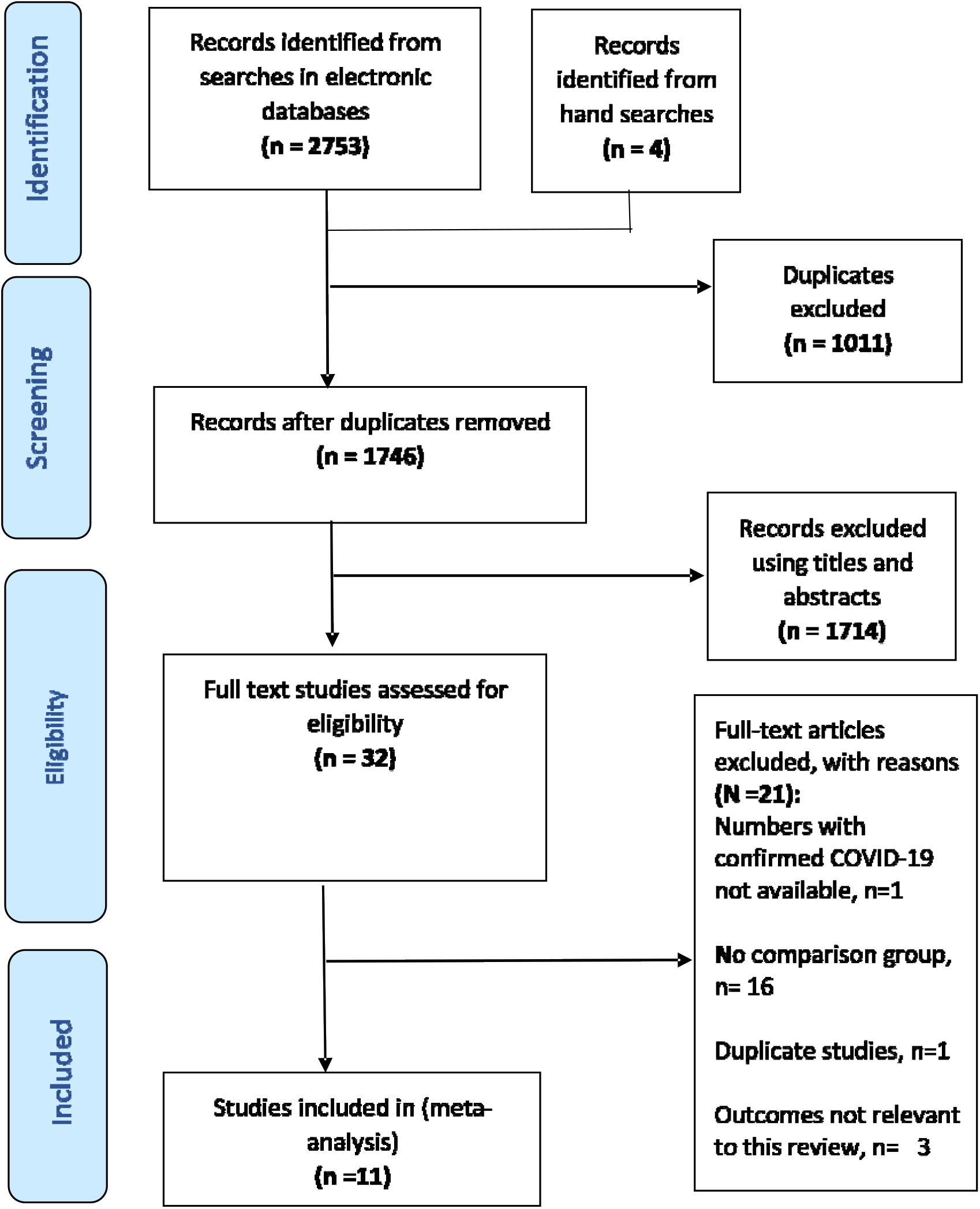
Flow chart for the systematic review and meta-analysis

### Characteristics of included studies

The included studies were from 4 countries and three regions only, namely, Europe (the United Kingdom and Spain), North America (the United States of America) and Sub-Saharan Africa (South Africa) (Fig 2). Six (17,26, 35–38) of the included studies were from the USA, followed by two studies (16, 39) from South Africa, one study (40) from the UK, and one study (41) from Spain (Fig 2). One study (42) included participants from both the USA and Spain.

**Fig.2.**
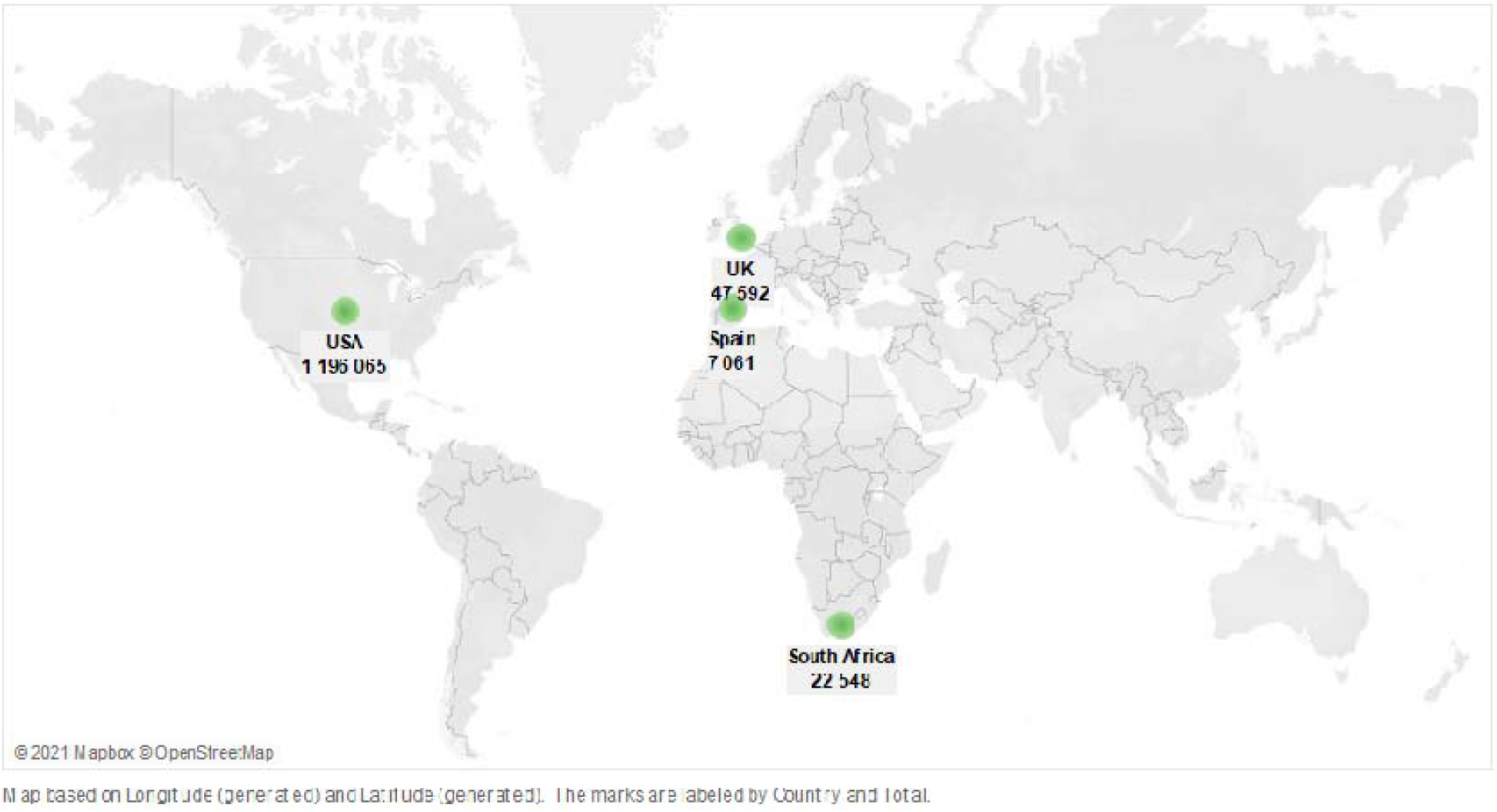
Map showing countries of origin of the included studies NB – The map shows the total participants with confirmed COVID-19 from each country

The total participants from all the included studies were 23 787 203, of which 668 797 were PLHIV and the remainder had no HIV. However, the total number of participants who had confirmed COVID-19 and were therefore assessed for the outcomes of this meta-analysis were 1 268 676 of which 13 886 were PLHIV and 1 254 790 were individuals without HIV. At the time of submission of this review, nine of the included studies (16,17,35–41) were published in peer reviewed journals. Of the remaining two studies, one was a letter to the editor (26) and the other a preprint (47). Four studies (16,17,26,42) did not report the mean or median ages of included participant but reported percentages in various age groups. Of the studies which reported the mean or median age of included participants, the smallest reported median age was 41.6 years in PLHIV in one study (31) and the oldest median age was 74 years, reported by Geretti et al. 2020, in individuals without HIV (40).

Of the included studies, there were five studies with small sample sizes which ranged from 63 participants to 493 participants. The remaining seven big studies had sample sizes ranging from 8 912 participants to 19 453 561 participants. The biggest of the included studies were population-based studies which compared mortality between PLHIV and individuals without HIV from the USA, Tesoriero et al. 2021 (19 453 561 participants) (17) and South Africa, Boulle et al (3 460 932 participants) (16). For these population-based studies we analyzed participants with confirmed COVID-19 only; from Tesoriero et al. 2020 (2988 PLHIV and 375 260 without HIV) (17) and from Boulle et al. 2020 (3978 PLHIV and 18330 without HIV) (16).

In terms of HIV-related characteristics, only one study reported data on the duration of HIV in PLHIV (48). Six studies (35, 37–41) reported data on the proportion of participants on ART which ranged from 88% to 100%. The proportion of participants with suppressed HIV viral load (lower than detectable limit (LDL)) ranged from 71% in South Africa (39) to 88% in the USA (17), with data reported from only five studies (18, 41–43, 45).

### Quality assessment of included studies

The MASTER score ranged from 7 to 17 out of a possible 36 (Supplementary Table 2). Due to their observational nature, the studies were mostly deficient in the domains of equal ascertainment, equal implementation, equal prognosis, and temporal precedence (Supplementary Fig 1). All the included studies were observational cohorts and did not blind caregivers, analysts and outcome assessors to participants HIV status or did not explicitly report doing so. Studies did not report whether COVID-19 treatment was delivered equally between PLHIV and individuals without HIV. In all the studies, it was also not clear if co-interventions that could impact the outcome were comparable between PLHIV and participants without HIV. Most of the studies, except the matched studies (14, 41, 42, 43), did not have design features in place to account for confounding.

### Mortality in PLHIV compared to individuals without HIV

Ten studies (16,17,26, 35–41), with a total sample size of 510 614 of which 7 836 were PLHIV and 502 778 were individuals without HIV, reported data on COVID-19 related mortality. Three studies (17, 18, 46), from South Africa, the UK and the USA, which contributed more than three-quarters of the total combined participants in this review, found a higher risk for mortality in PLHIV with strong evidence against the model hypothesis, after adjusting for confounders, with effect sizes ranging from an adjusted standardized rate ratio of 1.2 (95%CI 1.1-1.4) to an adjusted hazard ratio of 2.1 (95%CI 2.0-3.4). All the remaining studies reported no clinically important differences in mortality by HIV status (14, 28, 41, 43, 45, 48). Apart from Hadi et al. 2020 (14), all these remaining studies either had small sample sizes, overall or small numbers of PLHIV (Table 2).

**Table 2.**
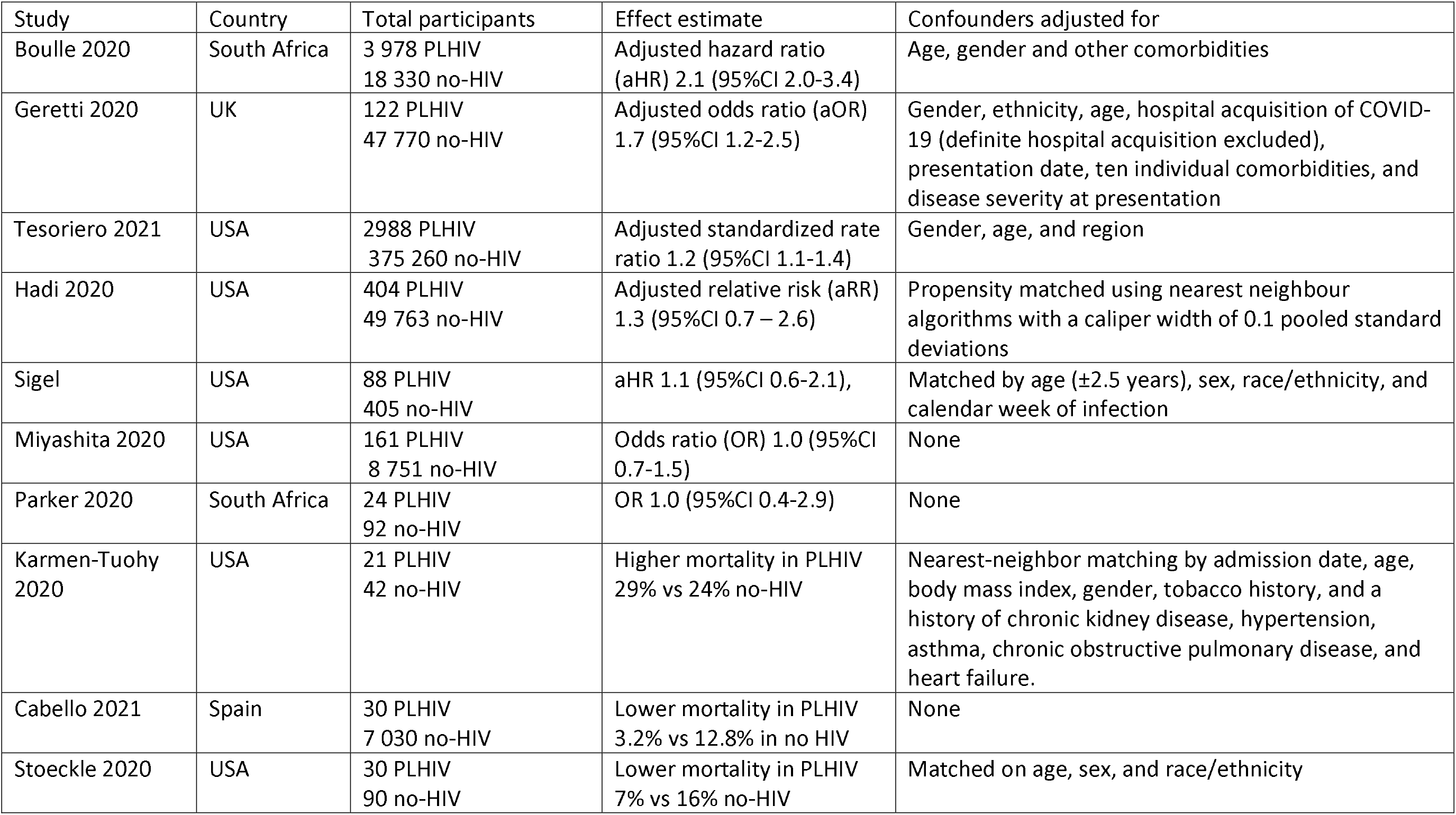
HIV and risk of mortality from COVID-19

| Study | Country | Total participants | Effect estimate | Confounders adjusted for |
| --- | --- | --- | --- | --- |
| Boulle 2020 | South Africa | 3 978 PLHIV<br>18 330 no-HIV | Adjusted hazard ratio (aHR) 2.1 (95%CI 2.0-3.4) | Age, gender and other comorbidities |
| Geretti 2020 | UK | 122 PLHIV<br>47 770 no-HIV | Adjusted odds ratio (aOR) 1.7 (95%CI 1.2-2.5) | Gender, ethnicity, age, hospital acquisition of COVID-19 (definite hospital acquisition excluded), presentation date, ten individual comorbidities, and disease severity at presentation |
| Tesoriero 2021 | USA | 2988 PLHIV<br>375 260 no-HIV | Adjusted standardized rate ratio 1.2 (95%CI 1.1-1.4) | Gender, age, and region |
| Hadi 2020 | USA | 404 PLHIV<br>49 763 no-HIV | Adjusted relative risk (aRR) 1.3 (95%CI 0.7 – 2.6) | Propensity matched using nearest neighbour algorithms with a caliper width of 0.1 pooled standard deviations |
| Sigel | USA | 88 PLHIV<br>405 no-HIV | aHR 1.1 (95%CI 0.6-2.1), | Matched by age ($\pm 2.5$ years), sex, race/ethnicity, and calendar week of infection |
| Miyashita 2020 | USA | 161 PLHIV<br>8 751 no-HIV | Odds ratio (OR) 1.0 (95%CI 0.7-1.5) | None |
| Parker 2020 | South Africa | 24 PLHIV<br>92 no-HIV | OR 1.0 (95%CI 0.4-2.9) | None |
| Karmen-Tuohy 2020 | USA | 21 PLHIV<br>42 no-HIV | Higher mortality in PLHIV 29% vs 24% no-HIV | Nearest-neighbor matching by admission date, age, body mass index, gender, tobacco history, and a history of chronic kidney disease, hypertension, asthma, chronic obstructive pulmonary disease, and heart failure. |
| Cabello 2021 | Spain | 30 PLHIV<br>7 030 no-HIV | Lower mortality in PLHIV 3.2% vs 12.8% in no HIV | None |
| Stoeckle 2020 | USA | 30 PLHIV<br>90 no-HIV | Lower mortality in PLHIV 7% vs 16% no-HIV | Matched on age, sex, and race/ethnicity |

In pooled analyses, there were a total of 428 deaths out of 7 836 PLHIV, compared to 33 440 deaths out of 502 778 individuals without HIV. The pooled unadjusted OR for overall mortality in PLHIV, compared to individuals without HIV, was 1.3 (95%CI 0.9-2.0, p =0.198) with high heterogeneity (I^2^ = 78.6%, p<0.001) (Fig. 3). The Doi plot (Supplementary Fig. 2) showed major asymmetry, indicative of possible publication bias. With the two South African studies removed, subgroup analysis of the European and North American studies did not change the OR for mortality (OR 1.4, 95%CI 0.8 – 2.6, p = 0.219), but the heterogeneity remained (I^2^ =77.1%, p <0.001) (Supplementary Fig. 3). Data from the two South African studies showed no clinically important differences in overall mortality between PLHIV and individuals without HIV with unadjusted ORs of 0.9 (95%CI 0.3 – 2.5) in one study (39) and 1.04 (95%CI 0.9 – 1.3) in the other (16) (Fig. 3).

**Fig. 3.**
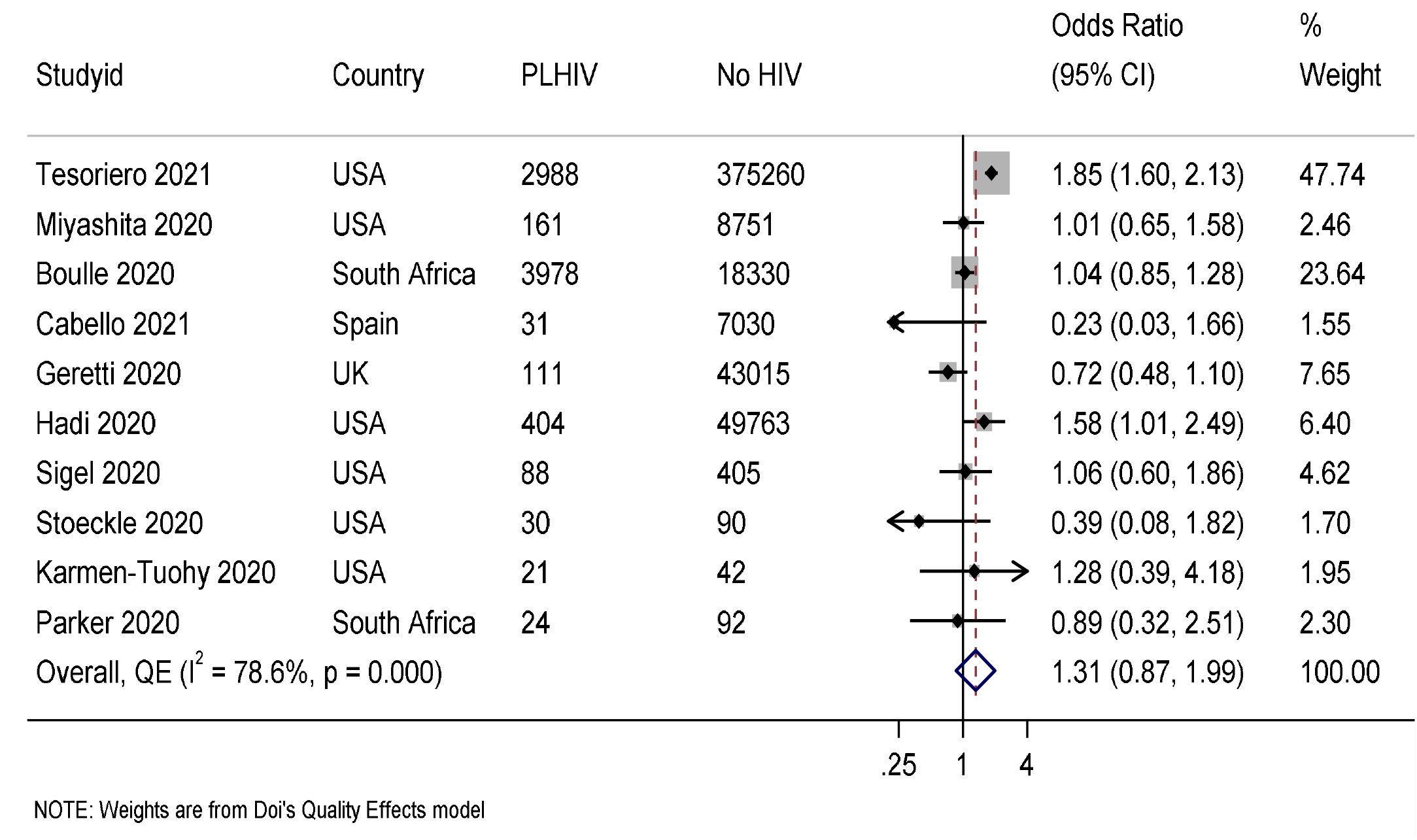
Odds ratios for overall mortality in PLHIV compared to individuals without HIV \*\*\**Columns headed “PLHIV” and “no HIV” depict total numbers of indiduals assessed for the outcome* Only three studies (16,17,26) reported data on mortality in age groups, with a total of 7 127 PLHIV and 402 341 individuals without HIV. In people aged <60 years, the pooled odds of mortality in PLHIV were almost three-fold the odds for mortality in individuals without HIV (OR 2.7, 95%CI 1.1-6.5, I^2^ =95.7%, p < 0.001). In people aged ≥60 years, the odds for mortality in PLHIV were similar to the odds in the overall model (OR 1.3, 95%CI 0.8 – 2.0, I^2^ = 58.2%, p = 0.092) (Fig. 4).

**Fig. 4.**
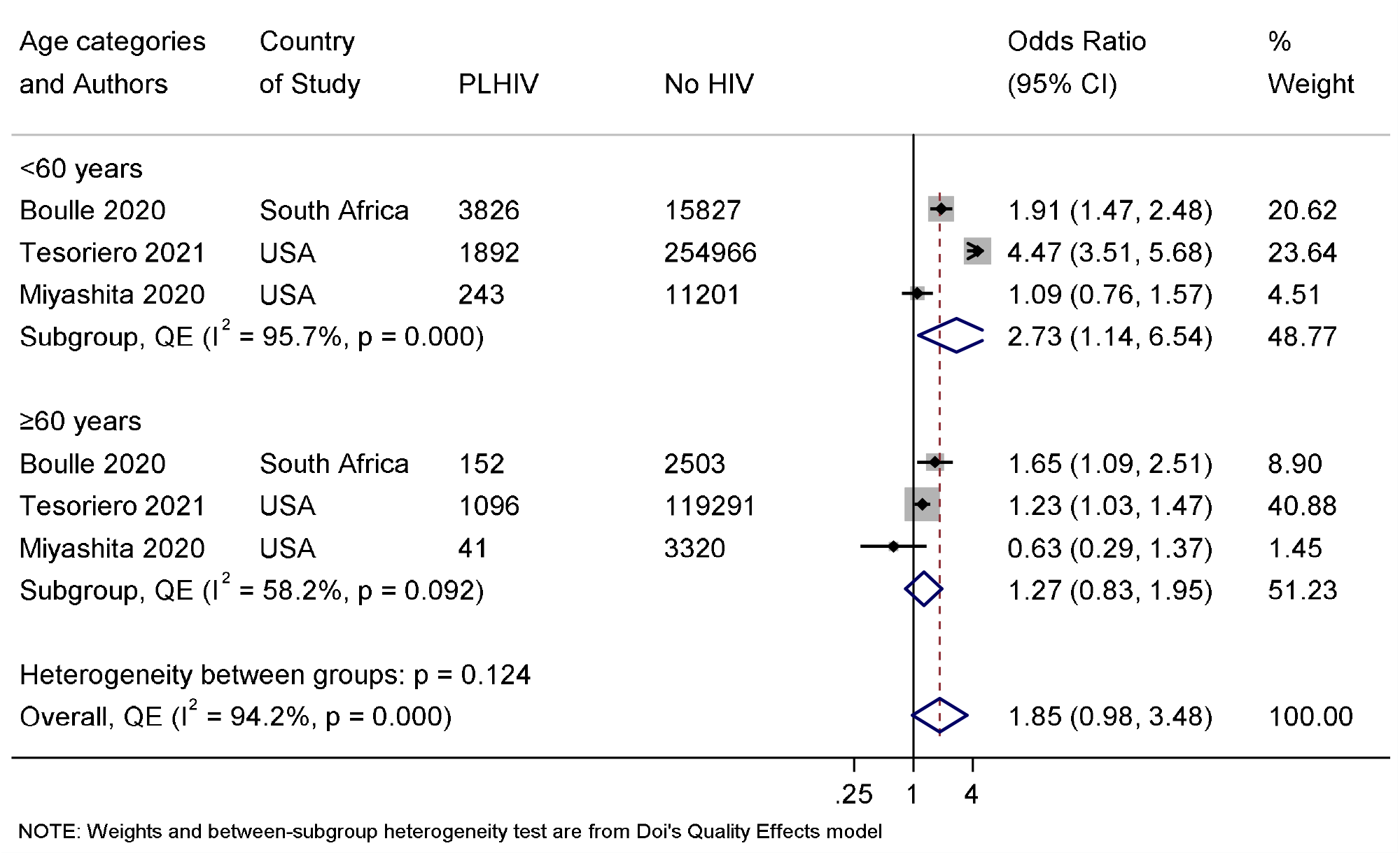
Odds ratio for mortality in PLHIV compared to individuals without HIV, by age group ***Columns headed “PLHIV” and “no HIV” depict total numbers of individuals assessed for the outcome **Data for Miyashita et al. 2021 used the 66 years age group cut-off

### Hospitalization for COVID-19 in PLHIV compared to individuals without HIV

Five studies (16,17,36,41,42) with a total sample size of 1 215 846 of which 13 451 were PLHIV and 1 202 395 had no HIV, reported data on hospitalization due to COVID-19. Adjusted analyses from two studies (17, 36) from the USA, showed a higher risk of hospitalization in PLHIV with a standardized relative risk (SRR ) of 1.4 (95%CI 1.3-1.5) and an adjusted relative risk (aRR) 1.7 (95% 1.2 – 2.4) respectively. Of the remaining three studies with unadjusted effect measures, two studies showed a higher risk of hospitalization in PLHIV while one showed a lower risk of hospitalization in PLHIV compared to individuals without HIV (Table 3).

**Table 3.** HIV and the risk of hospitalization from COVID-19

| Study | Country | Total participants | Effect estimate | Confounders adjusted for |
| --- | --- | --- | --- | --- |
| Tesoriero 2021 | USA | 2988 PLHIV<br>375 260 no-HIV | Hospitalization –<br>standardized rate ratio<br>(SRR) 1.4 (95%CI 1.3-1.5), | Gender, age, and region |
| Hadi 2020 | USA | 404 PLHIV<br>49 763 no-HIV | Adjusted relative risk-<br>aRR 1.7 (95% 1.2 – 2.4) | Propensity matched<br>using nearest neighbour<br>algorithms with a<br>caliper width of 0.1<br>pooled standard<br>deviations |
| Boulle 2020 | South Africa | 3 978 PLHIV<br>18 330 no-HIV | OR 1.2 (95%CI 1.1 –<br>1.3) | None |
| Kohler 2020 | USA & Spain | 6050 PLHIV<br>752 012 no-HIV | Hospitalization (higher in<br>PLHIV 37.8% vs 27.8%) | None |
| Cabello 2021 | Spain | 30 PLHIV<br>7 030 no-HIV | Hospitalization (lower in<br>PLHIV 48.4% vs 57.7%) | None |

In pooled analyses, a total of 3 583 individuals out of 13 451 PLHIV were hospitalized for COVID-19, compared to 233 051 individuals out of 1 202 395 individuals without HIV. The pooled OR for hospitalization in PLHIV, compared to individuals without HIV was 1.7 (95% CI 1.3-2.1,) with high heterogeneity (I^2^ = 96.0%, p<0.001) (Fig. 5). The Doi plot (Supplementary Fig. 4) showed major asymmetry, indicative of possible publication bias. We repeated the analysis with the European and US studies only, but the OR estimates did not change significantly (OR 1.8, 95% 1.5 – 2.2) and the high heterogeneity remained (I^2^ = 87.0%, p<0.01) (Supplementary Fig 5).

**Fig. 5.**
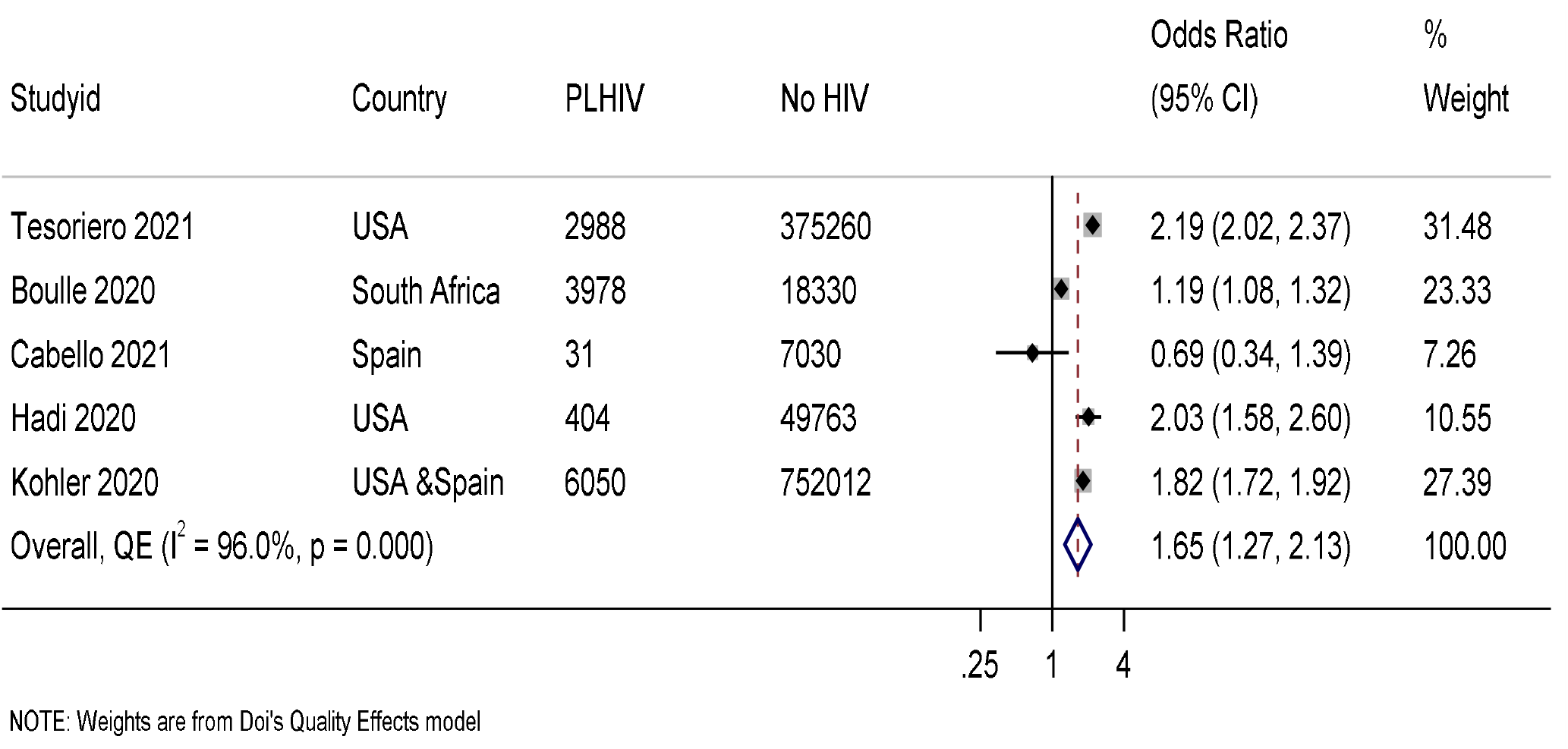
Odds ratio for hospitalization for COVID-19 in PLHIV compared to individuals without HIV \*\*\**Columns headed “PLHIV” and “no HIV” depict total numbers of individuals assessed for the outcome* Only one study (17) reported data on hospitalization in different age groups, so it was not possible to carry out subgroup meta-analysis. Similar to the pattern observed for mortality, the OR for hospitalization in PLHIV, compared to those without HIV appeared to be highest in people aged <60 years (OR 2.8, 95%CI 2.5-3.2), and lower in people aged ≥60 years (OR 1.7, 95%CI 1.5-1.9) (Supplementary Fig 6).

### Intensive care services for COVID-19 in PLHIV compared to individuals without HIV

Eight studies (26,35, 37–41) with a total of 6 239 PLHIV and 693 066 individuals without HIV, reported data comparing the need for intensive care services. The three matched studies showed differing results, with one showing a higher risk of intensive care services, another showing no differences in proportions requiring mechanical ventilation and the third finding a lower risk of both ICU and mechanical ventilation in PLHIV. Unadjusted analysis of data from Geretti et al. showed an almost three-fold increase in odds of intensive care services in PLHIV compared to those without HIV. The results of the other studies that carried out unadjusted analysis were unequivocal (Table 3).

**Table 3.**
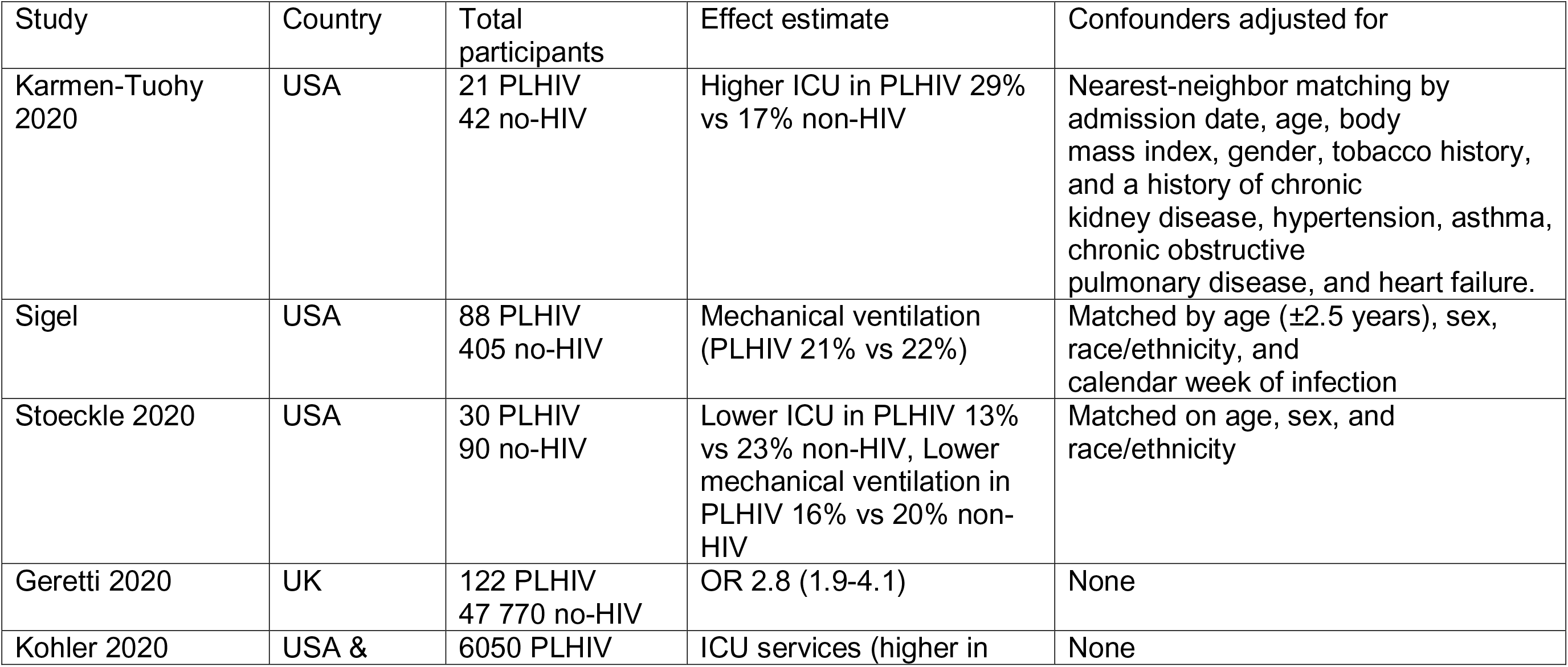
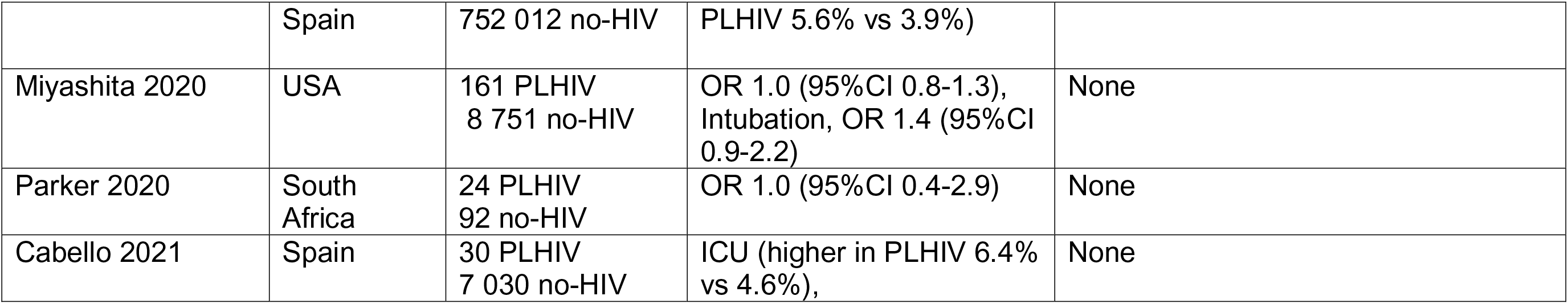
HIV and the need for intensive care services due to COVID-19

In pooled analysis, 401 individuals out of 6 239 PLHIV needed intensive care services, compared to 29 006 individuals out of 693 066 individuals without HIV. The pooled OR for the need of intensive care services for PLHIV, compared to individuals without HIV, was 1.4 (95% CI 0.9-2.0) with substantial heterogeneity (I^2^ = 74%, p <0.001 (Fig. 6). The Doi plot (Supplementary Fig. 7) showed major asymmetry, suggesting possible publication bias.

**Fig. 6.**
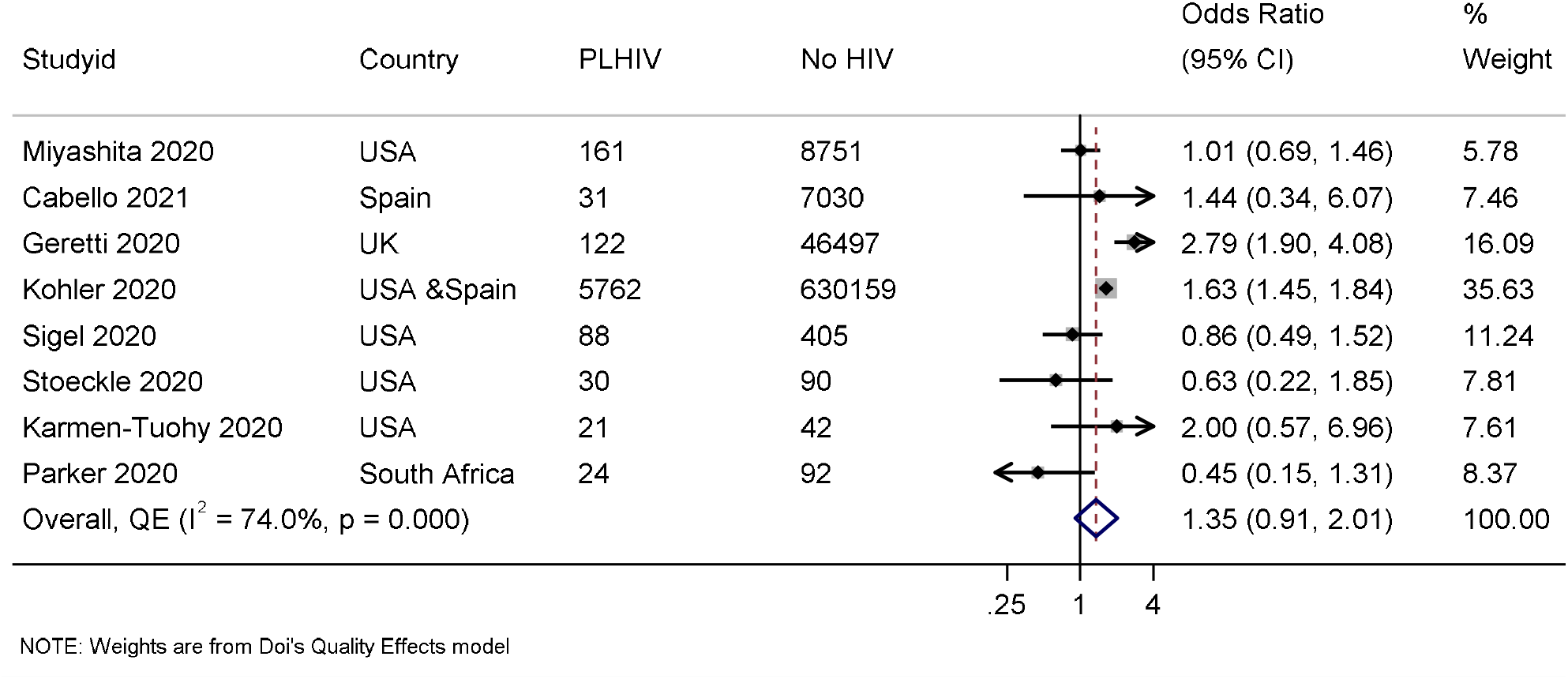
Odds ratio for intensive care services for COVID-19 in PLHIV compared to individuals without HIV \*\*\**Columns headed “PLHIV” and “no HIV” depict total numbers of individuals assessed for the outcome* Only one study (26) reported data on the risk for intensive care services by age groups. Similar patterns observed for mortality and hospitalization were observed with intensive care services in this study, with a higher OR for intensive care services for PLHIV in the <66 years age group (OR 1.2, 95%CI 0.9 – 1.6), and a lower OR in those aged ≥66 years (unadjusted OR 0.4, 95%CI 0.2 – 1.0) (Supplementary Fig. 8).

### Comparison of the prevalence chronic comorbidities individuals with COVID-19 by HIV status

Six of the included studies (16,17,26,36,39,40) with a total of 132 495 individuals of which 4 909 were PLHIV and 127586 with no HIV, reported data on comorbidities by HIV status in individuals with COVID-19 (Supplementary Fig 9). From four studies, the prevalence of hypertension was higher in PLHIV in two studies from the USA (26, 36) with ORs of 2.0 and 2.2, respectively, with strong evidence against the null hypothesis in both studies. However, data from two studies from South Africa (16, 39) suggested a lower prevalence of hypertension in PLHIV, with ORs of 0.7 and 0.9, respectively, but with weak evidence against the model hypothesis at their sample sizes. The pooled odds ratio suggested no difference in the prevalence of hypertension by HIV status (OR 0.9, 95%CI 0.3 – 2.9) with high between-study heterogeneity (I^2^ =99.5%).

Five studies provided data on proportions of cases with diabetes by HIV status. Data from two studies from the USA (26, 36) suggested a higher prevalence of diabetes in PLHIV with ORs of 1.7 and 1.6, respectively, and strong evidence against the model hypothesis while data from one study from South Africa (39) also suggested a higher prevalence of diabetes in PLHIV but with weak evidence against the model hypothesis at their sample size. However, data from two studies suggested a lower prevalence of diabetes in PLHIV, one from South Africa (16) with an OR of 0.6 and strong evidence against the model hypothesis, and the other from the UK (40), but with weak evidence against the model hypothesis at their sample size. The pooled OR of 0.8 (95%CI 0.3 – 1.8, I^2^ = 98.5%), suggested no difference in the prevalence of diabetes by HIV status in the included studies, again with high heterogeneity.

Four studies provided data on CVD, with a higher prevalence of CVD in PLHIV from two from the USA, one study (36) with an OR of 1.8 and strong evidence against the model hypothesis and another (26) with an OR of 1.3 and weak evidence against the model hypothesis. Data from the remaining two studies suggested a lower prevalence of CVD in PLHIV, with an OR of 0.4 and strong evidence against the model hypothesis from one study from the UK (40) and an OR of 0.8 and weak evidence against the model hypothesis from the other study from South Africa (39). The pooled OR suggested no difference in the prevalence of CVD by HIV status, with high heterogeneity between the studies (OR 1.2, 95%CI 0.4 – 3.1, I^2^ = 96.8%).

Four studies provided data on cases with chronic kidney disease by HIV status, with data from two studies, one from South Africa (39) and one from the USA (26) suggesting a higher prevalence of chronic kidney disease in PLHIV with ORs 7.8 and 2.9, respectively, both with strong evidence against the model hypothesis. However, data from the other two studies suggested a lower prevalence of chronic kidney disease in PLHIV with an OR of 0.2 and strong evidence against the model hypothesis in one study from the UK (40) and an OR of 0.9 and weak evidence against the model hypothesis in the other study from South Africa (16). The pooled odds suggested no clinically important difference in the prevalence of chronic kidney disease by HIV status, with high heterogeneity (OR 0.8, 95%CI 0.8-1.2, I^2^= 97.6%).

Four studies provided data on asthma, with data from two studies, one from the USA (36) and another from South Africa (39) suggesting a higher prevalence of asthma in PLHIV, with ORs of 1.8 and 2.0 and strong evidence against the model hypothesis in both. Data from the remaining two studies suggested a lower prevalence of asthma in PLHIV, with an OR of 0.7 and strong evidence against the model hypothesis from one study from the South Africa (16) and an OR of 0.7 and weak evidence against the model hypothesis from the other study from the UK (40). The pooled OR suggested no difference in the prevalence of asthma by HIV status, with high heterogeneity between the studies (OR 1.0, 95%CI 0.4 – 2.1, I^2^ = 93.8%).

Only three studies provided data on the prevalence of obesity with data from two studies (36, 40)suggesting a higher prevalence of obesity in PLHIV, with ORs of 1.3 and 1.6, respectively, and strong evidence against the model hypothesis, and data the remaining study from South Africa (39) suggested a lower prevalence but with weak evidence against the model hypothesis at their sample size.

Two studies reported data on the prevalence of malignancies, with a high prevalence in PLHIV from one study from South Africa (39) (OR 1.2, 95%CI 0.0-31.5) and a lower prevalence in PLHIV from the other study from the UK (40) (OR 0.3, 95%CI 0.1 – 0.8) but with weak evidence against the model hypothesis in both studies. Two studies reported data on dyslipidaemia, with a high prevalence of dyslipidaemia and strong evidence against the model hypothesis in PLHIV in one study from the USA (26) (OR 2.3, 95%CI 1.6 -3.1) and a lower prevalence but weak evidence against the model hypothesis in the other study from South Africa (OR 0.4, 95%CI 0.0 – 3.3) (Supplementary Fig 9).

## Discussion

In this systematic review and meta-analysis of 1 273 266 individuals from 11 studies from North America, Europe and sub-Saharan Africa, we compared mortality, hospitalization and intensive care services between individuals with and without HIV. Overall, compared to individuals without HIV, the estimated effect of HIV on either mortality or intensive care services suggested some worsening with very weak evidence against the model hypothesis at this sample size. However, in individuals aged <60 years, the estimated odds of mortality were increased by 2.7-fold in PLHIV with strong evidence against the model hypothesis at this sample size. HIV was also associated with an estimated 1.7-fold increase in odds of hospitalization for COVID-19 for PLHIV that suggested worsening, also with strong evidence against the model hypothesis at this sample size. Lastly, we found no qualitative differences in the proportion of individuals with chronic comorbidities between PLHIV and individuals without HIV.

Findings from several existing systematic reviews and primary studies are conflicting. Three existing systematic review (4,7,8) suggested no difference in COVID-19 severity between PLHIV and individuals without HIV, although their analyses were narrative. Two of the studies that we included, a letter to the editor with unadjusted analysis (26) and a matched cohort study with 63 participants (38) also reported no differences in the risk of death by HIV status. However, other studies have reported a higher risk of mortality from COVID-19 for PLHIV. One recent systematic review reported a higher risk of mortality in PLHIV with a risk ratio 1.8 (95%CI 1.2-2.6) (19), although they included four studies only, including data from Bhaskaran et al. 2020 (18), where totals with confirmed COVID-19 were not available. The large population-based cohort studies which we included all showed a higher risk of death from COVID-19 in PLHIV after adjusting for confounders. In adjusted models, the estimated effects of HIV on mortality from the large cohorts ranged from a standardized rate ratio of 1.2 (17) in the USA to a hazard ratio of around two from South Africa and the UK (17, 46). Notably the pooled OR is not adjusted for confounders as the included studies reported different effect measures.

Although in the overall analysis the increase in the risk of mortality was marginal, in subgroup analysis, PLHIV aged <60 years had 2.7 times the odds of mortality from COVID-19 compared to similar aged individuals without HIV. This is a pertinent finding, especially for countries in sub-Saharan African where most PLHIV are resident, as they are in the young adult and middle-aged adult age groups. It is not yet clear why PLHIV may have a higher risk for mortality from COVID-19 as there is currently a lack of consensus about the mechanisms that may increase the risk of mortality in PLHIV. Chronic inflammation and immune deficiency due to the HIV disease process may be key contributors to the higher susceptibility for severe COVID-19. Despite adequate antiretroviral therapy (ART), PLHIV may still have persistent immune dysregulation which might increase the susceptibility and severity of COVID-19 in this cohort (43, 44). On the other hand, the same dysfunctional immune responses may offer protection against the cytokine storm triggered in severe COVID-19 (45). A number of antiretroviral drugs have also shown in vitro activity against SARS-CoV-2 (46), but whether this confers protection from severe COVID-19 in individuals on ART remains unclear.

Our pooled OR suggests that PLHIV, compared to individuals without HIV, have a 1.6-fold increase in the odds of being hospitalized and a 1.4-fold increase in odds for intensive care service for COVID, although with weak evidence against the model hypothesis for the later. Our findings are in agreement with most of the studies which we included, except three studies which reported lower hospitalization rates in PLHIV (41), and lower intensive care services in PLHIV (35, 37). Three (17,36,42) of the included studies reported higher risk of hospitalization in PLHIV, and another four reported a higher risk of intensive care services in PLHIV (26,38,41,42). Again, these findings suggest a higher risk for severe disease and higher utilization of health resources in PLHIV and have implications for high-burden HIV countries in Sub-Saharan Africa such as South Africa. If COVID-19 is not brought under control and case counts continue increasing, these countries may experience higher levels of hospitalizations and need for intensive care services. This is likely to be worsened by the high prevalence of undiagnosed chronic comorbidities such as diabetes and hypertension (47). Data from the included studies that comorbidities were likely to occur equally between the two groups and thus are not a major influence on the association we report. In studies of people without COVID-19, HIV is associated with a higher risk of cardiovascular disease (48) and ART has been associated with metabolic syndrome (49) and high fasting glucose (50). However, the evidence is not conclusive as other meta-analyses have found no associations between ART and diabetes (51) and hypertension (52). Although the influence of comorbid conditions cannot be ruled out, our findings suggest that the mechanism for the more severe COVID19 disease course in PLHIV may lie with the HIV disease process.

Although good progress had been made toward reaching the 90-90-90 (53) targets, however the targets are not yet met globally as only 59% of PLHIV had viral suppression in 2019 (5). The COVID-19 pandemic has disrupted the HIV care cascade especially in Sub-Saharan Africa (54), and may result in higher numbers of PLHIV with uncontrolled HIV and more vulnerable to COVID-19. There is need to prioritize prevention, including prioritizing vaccine access, care and treatment of COVID-19 in PLHIV.

Our study has several limitations. The studies included in this synthesis are observational studies and any inferences about the strength of the relationship between HIV and severe COVID-19 need to be made cautiously. Further, confounding variables, such as age and comorbidities, and type of HIV treatment may have affected some of the findings from included studies. It was not possible to do a meta-analysis of adjusted effect sizes as the studies reported different effect measures. We were also not able to carry out subgroup analysis on some of these possible confounders because of lack of data from included studies. Due to a lack of data from included studies, we were not able to analyse the effect of being on treatment for HIV, viral load and CD4 counts on COVID-19 severity and mortality. However, it is worth noting that one study found that the type of anti-retroviral treatment had no beneficial impact on COVID-19 severity or risk of death (13). A strength of this study is that it was carried out rigorously using the updated PRISMA guidelines for systematic reviews and meta-analyses. Further, this meta-analysis included studies where participants had confirmed COVID-19 only and used quality adjusted effects model to take into consideration the study quality of included studies, and therefore the risk estimates we reported are adjusted for the variations in study quality of the included observational studies.

## Conclusion

PLHIV may be at a higher risk of death and hospitalisation from COVID-19, compared to individuals without HIV. The differential mortality between individuals with and without HIV is seen more in people aged <60 years and thus PLHIV should be considered at greater risk of severity than those without HIV in the population. The findings of our study have several implications considering the high global burden of HIV, especially in Sub-Saharan Africa (55). Around two-thirds of PLHIV are in the Sub-Saharan Africa region, which has health systems which are already overstretched and fragile. The COVID-19 pandemic worsened in the period December 2020-February 2021 but case counts and mortality have remained lower than in many other regions. South Africa has been the exception, with higher case counts and deaths than other countries in Sub-Saharan. However, the possibility that COVID-19 may worsen in the coming months remains open. Compounding the vulnerable health systems in Sub-Saharan Africa are several issues which include difficulties in carrying out COVID-19 preventive measures (56), lower pandemic preparedness and a lack of safety nets for the vulnerable in a pandemic (57). Further, Sub-Saharan Africa is lagging behind many other countries in COVID-19 vaccinations (58), and this may mean that subsequent COVID-19 infection waves are expected and may have severe effects.

## Data Availability

Data are available from the included studies

## Abbreviations

HIV: Human Immunodeficiency Virus PLHIV – People living with HIV.
COVID-19: Coronavirus disease of 2019
SARS-CoV-2: severe acute respiratory syndrome coronavirus 2
RNAdRNAp: RNA-dependent RNA polymerase
ICU: Intensive care unit
UK: United Kingdom
USA: United States of America
ART: Antiretroviral therapy
OR: Odds ratio
95%CI: 95% confidence interval
MASTER: MethodologicAl STandard for Epidemiological Research

## CRediT Authors’ contributions

LM- conceptualisation, data curation, formal analysis, investigation, visualisation, writing – original draft, and writing – review & editing

AA - data curation, visualisation, investigation, writing – review & editing

RA - data curation, visualisation, investigation, writing – review & editing

NI - data curation, investigation, writing – review & editing

AC - data curation, investigation, writing – review & editing

SAR - formal analysis, investigation, methodology, supervision, validation, visualisation, writing – review & editing

TC - conceptualisation, data curation, formal analysis, investigation, methodology, project administration, resources, software, supervision, validation, visualisation, writing – original draft, and writing – review & editing

