## Supplementary material for "The risk of severe COVID-19 and mortality from COVID-19 in people living with HIV compared to individuals without HIV - a systematic review and meta-analysis of 1 268 676 individuals": HIV COVID-19 Supplementary materials (from another version of this preprint: version 1, 2021-07-05)

#### Supplementary Table 1 – Pubmed Search strategy

| | Search stratergy |
|---|---|
| COVID-19 | ((((("covid 19"[Supplementary Concept] OR "covid 19"[All Fields]) OR "covid19"[All Fields]) OR (("coronavirus"[MeSH Terms] OR "coronavirus"[All Fields]) OR "coronaviruses"[All Fields])) OR ((("novel"[All Fields] OR "novel s"[All Fields]) OR "novels"[All Fields]) AND (("coronavirus"[MeSH Terms] OR "coronavirus"[All Fields]) OR "coronaviruses"[All Fields]))) OR (("severe acute respiratory syndrome coronavirus 2"[Supplementary Concept] OR "severe acute respiratory syndrome coronavirus 2"[All Fields]) OR "sars cov 2"[All Fields])) OR "COVID"[All Fields]) OR ((((((("covid 19"[All Fields] OR "covid 2019"[All Fields]) OR "severe acute respiratory syndrome coronavirus 2"[Supplementary Concept]) OR "severe acute respiratory syndrome coronavirus 2"[All Fields]) OR "2019 ncov"[All Fields]) OR "sars cov 2"[All Fields]) OR "2019ncov"[All Fields]) OR (("wuhan"[All Fields] AND ("coronavirus"[MeSH Terms] OR "coronavirus"[All Fields])) OR ("coronaviurs" [All Fields] AND "disease" [All Fields]) OR ("2019-nCoV" [All Fields] AND "acute" [All Fields] AND "respiratory" [All Fields] AND "disease" [All Fields]) OR ("Novel" [All Field] AND "coronavirus" [All Fields] AND "pneumonia" [All Fields]) OR ("Wuhan" [All Fields] AND "pneumonia" [All Fields]) OR ("SARS-Cov-2" [All Fields]) (2019/12/1:2019/12/31[Date - Publication] OR 2020/1/1:2020/11/30[Date - Publication]))) |
| HIV | "HIV"[All Fields] OR "Hiv"[All Fields] OR "hiv"[All Fields] OR "Aids"[All Fields] OR “human immunodeficiency virus "[MeSH Terms] OR “human immunodeficiency virus "[All Fields] OR "acquired |

| | |
|---|---|
| | immunodeficiency syndrome"[All Fields] OR "acquired immunodeficiency syndrome"[MeSH Terms] OR "HIV"[ MeSH Terms] OR "Hiv"[ MeSH Terms] OR "hiv"[ MeSH Terms] OR "ARV"[All Fields] OR "ARV"[ MeSH Terms] OR "arv"[All Fields] OR "arv"[ MeSH Terms] "antiretroviral"[All Fields] OR "antiretroviral"[ MeSH Terms] OR "controlled HIV"[All Fields] OR "controlled HIV"[ MeSH Terms] OR "uncontrolled HIV"[All Fields] OR "uncontrolled HIV"[ MeSH Terms] |

#### Supplementary Table 2 – Assessment of study quality

| Safeguard item | Tesoriero 2021 | Miyashita 2020 | Boulle 2020 | Cabello 2021 | Geretti 2020 | Hadi 2020 | Kohler 2020 | Sigel 2020 | Stoeckle 2020 | Karmen-Tuohy 2020 | Parker 2020 | Total |
|---|---|---|---|---|---|---|---|---|---|---|---|---|
| 1. Data collected after the start of the study was not used to exclude participants or to select them into the analysis | 1 | 1 | 1 | 1 | 1 | 1 | 1 | 1 | 1 | 1 | 1 | 12 |
| 2. Participants in all comparison groups met the same eligibility requirements and were from the same population and timeframe | 1 | 0 | 1 | 1 | 1 | 1 | 0 | 1 | 1 | 1 | 1 | 10 |
| 3. Determination of eligibility and assignment to treatment group/exposure strategy was synchronized | 0 | 0 | 0 | 0 | 0 | 0 | 0 | 0 | 0 | 0 | 0 | 0 |
| 4. None of the eligibility criteria were common effects of exposure and outcome | 1 | 0 | 1 | 1 | 1 | 1 | 1 | 1 | 1 | 1 | 1 | 11 |
| 5. Any attrition (or exclusions after entry) is less than 20% of total participant numbers | 1 | 0 | 1 | 1 | 1 | 1 | 0 | 1 | 1 | 1 | 1 | 10 |
| 6. Missing data is less than 20% | 1 | 0 | 1 | 1 | 1 | 1 | 0 | 1 | 1 | 1 | 1 | 10 |
| 7. Analysis accounted for missing data | 1 | 0 | 1 | 1 | 1 | 0 | 0 | 1 | 1 | 1 | 1 | 9 |
| 8. Exposure variations/treatment deviations were less than 20% | 0 | 0 | 0 | 0 | 0 | 0 | 0 | 0 | 0 | 0 | 0 | 0 |
| 9. Variations in exposure or withdrawls | 0 | 0 | 0 | 0 | 0 | 0 | 0 | 0 | 0 | 0 | 0 | 0 |

| | | | | | | | | | | | | |
|---|---|---|---|---|---|---|---|---|---|---|---|---|
| after the start of the study were addressed by analysis | | | | | | | | | | | | |
| 10. Procedures for data collection of covariates were reliable and the same for all participants | 1 | 0 | 1 | 1 | 1 | 1 | 1 | 1 | 1 | 1 | 1 | 11 |
| 11. The outcome was objectively defined and/or reliably measured | 1 | 1 | 1 | 1 | 1 | 1 | 1 | 1 | 1 | 1 | 1 | 12 |
| 12. Exposures/ interventions were objectively defined and/ or reliably measured | 0 | 0 | 0 | 0 | 0 | 0 | 0 | 0 | 0 | 0 | 1 | 1 |
| 13. Outcome assessor(s) were blinded | 0 | 0 | 0 | 0 | 0 | 0 | 0 | 0 | 0 | 0 | 0 | 0 |
| 14. Participants were blinded | 0 | 0 | 0 | 0 | 0 | 0 | 0 | 0 | 0 | 0 | 0 | 0 |
| 15. Caregivers were blinded | 0 | 0 | 0 | 0 | 0 | 0 | 0 | 0 | 0 | 0 | 0 | 0 |
| 16. Analyst was blinded | 0 | 0 | 0 | 0 | 0 | 0 | 0 | 0 | 0 | 0 | 0 | 0 |
| 17. Care was delivered equally to all participants | 0 | 0 | 0 | 0 | 0 | 0 | 0 | 0 | 0 | 0 | 0 | 0 |
| 18. Cointerventions that could impact the outcome were comparable between groups or avoided | 0 | 0 | 0 | 0 | 0 | 0 | 0 | 0 | 0 | 0 | 0 | 0 |
| 19. Control and active interventions/ exposures are sufficiently distinct | 0 | 0 | 0 | 0 | 0 | 0 | 0 | 0 | 0 | 0 | 0 | 0 |
| 20. Exposure/intervention definition consistently applied to all participants | 1 | 1 | 1 | 1 | 1 | 1 | 1 | 1 | 1 | 1 | 1 | 12 |
| 21. Outcome definition consistently applied to all participants | 1 | 1 | 1 | 1 | 1 | 1 | 1 | 1 | 1 | 1 | 1 | 12 |
| 22. The time period between exposure and outcome is similar across patients and between groups or the analyses adjust for different lengths of follow-up of patients | 0 | 0 | 0 | 0 | 0 | 0 | 0 | 0 | 0 | 0 | 0 | 0 |
| 23. Design and/or analytic strategies were in place that addressed potential confounding | 0 | 0 | 0 | 0 | 1 | 1 | 0 | 1 | 0 | 0 | 0 | 3 |
| 24. Key confounders addressed through design or analysis were not common effects of exposure and outcome | 1 | 0 | 1 | 0 | 1 | 1 | 0 | 1 | 0 | 0 | 0 | 5 |
| 25. Key baseline characteristics / prognostic indicators for the study were | 0 | 0 | 0 | 0 | 0 | 0 | 0 | 0 | 0 | 0 | 0 | 0 |

| | | | | | | | | | | | | |
|---|---|---|---|---|---|---|---|---|---|---|---|---|
| comparable across groups | | | | | | | | | | | | |
| 26. Participants were randomly allocated to groups with adequate randomisation process | 0 | 0 | 0 | 0 | 0 | 0 | 0 | 0 | 0 | 0 | 0 | 0 |
| 27. Allocation procedure was adequately concealed | 0 | 0 | 0 | 0 | 0 | 0 | 0 | 0 | 0 | 0 | 0 | 0 |
| 28. Conflict of interests were declared and absent | 1 | 0 | 1 | 1 | 1 | 1 | 0 | 1 | 1 | 1 | 1 | 10 |
| 29. Analytic method was justified by study design or data requirements | 0 | 0 | 0 | 0 | 1 | 1 | 0 | 0 | 0 | 0 | 0 | 2 |
| 30. Computation errors or contradictions were absent | 1 | 1 | 1 | 1 | 1 | 1 | 0 | 1 | 1 | 1 | 1 | 11 |
| 31. There was no discernable data dredging or selective reporting of the outcome | 1 | 0 | 1 | 1 | 1 | 1 | 0 | 1 | 1 | 1 | 1 | 10 |
| 32. All subjects were selected prior to intervention/exposure and evaluated prospectively | 0 | 0 | 0 | 0 | 0 | 0 | 0 | 0 | 0 | 0 | 0 | 0 |
| 33. Carry-over or refractory effects were avoided or considered in the design of the study or were not relevant | 1 | 1 | 1 | 1 | 1 | 1 | 1 | 1 | 1 | 1 | 1 | 12 |
| 34. The intervention/ exposure period was long enough to have influenced the study outcome | 0 | 0 | 0 | 0 | 0 | 0 | 0 | 0 | 0 | 0 | 0 | 0 |
| 35. Dose of intervention/ exposure was sufficient to influence the outcome | 0 | 0 | 0 | 0 | 0 | 0 | 0 | 0 | 0 | 0 | 0 | 0 |
| 36. Length of follow-up was not too long or too short in relation to the outcome assessment | 0 | 0 | 0 | 0 | 0 | 0 | 0 | 0 | 0 | 0 | 0 | 0 |
| **Summary count of safeguard items** | **15** | **6** | **15** | **14** | **17** | **16** | **7** | **16** | **14** | **14** | **15** | |

#### Supplementary Fig 1 – Assessment of study quality

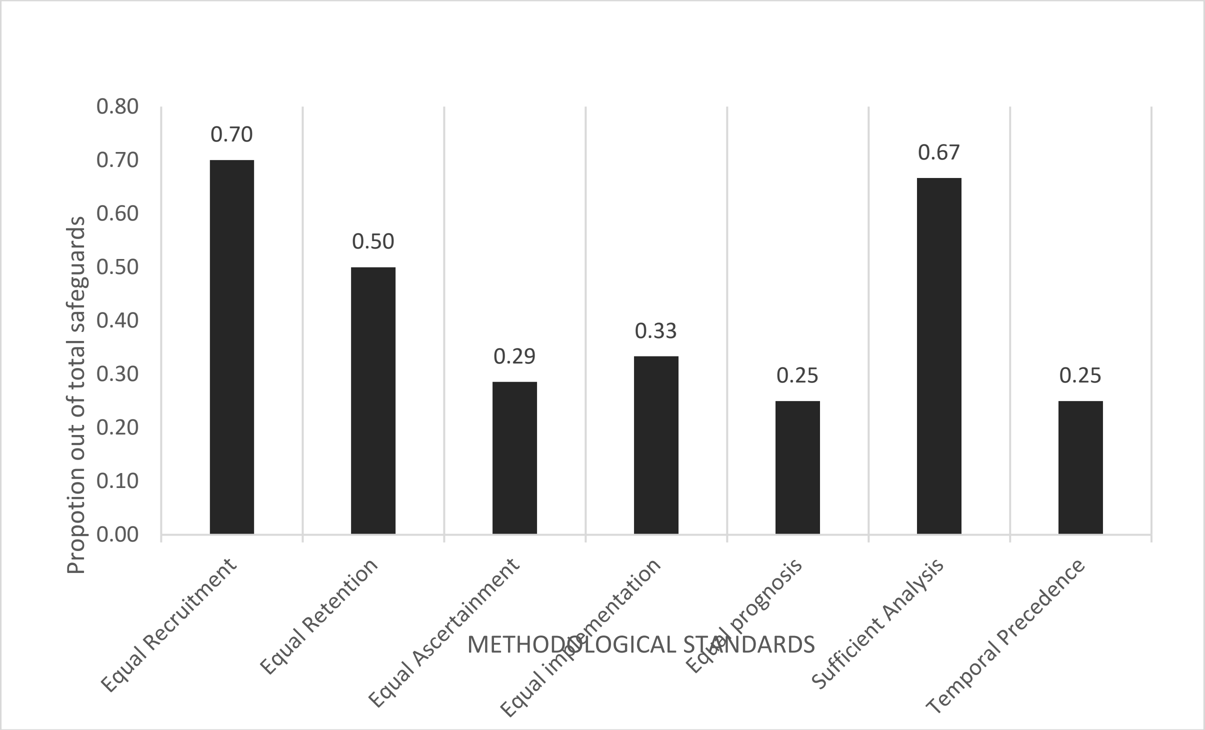


#### Supplementary Fig 2- Doi plot and funnel plot for mortality

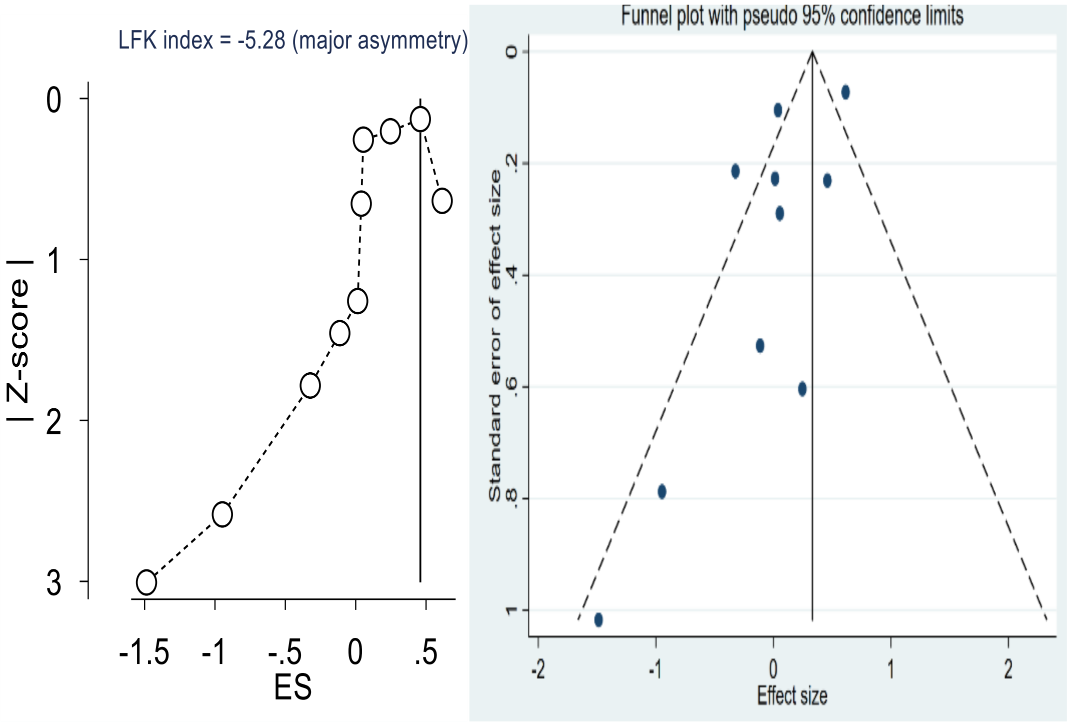

#### Supplementary Fig 3- Mortality without the South Africa studies

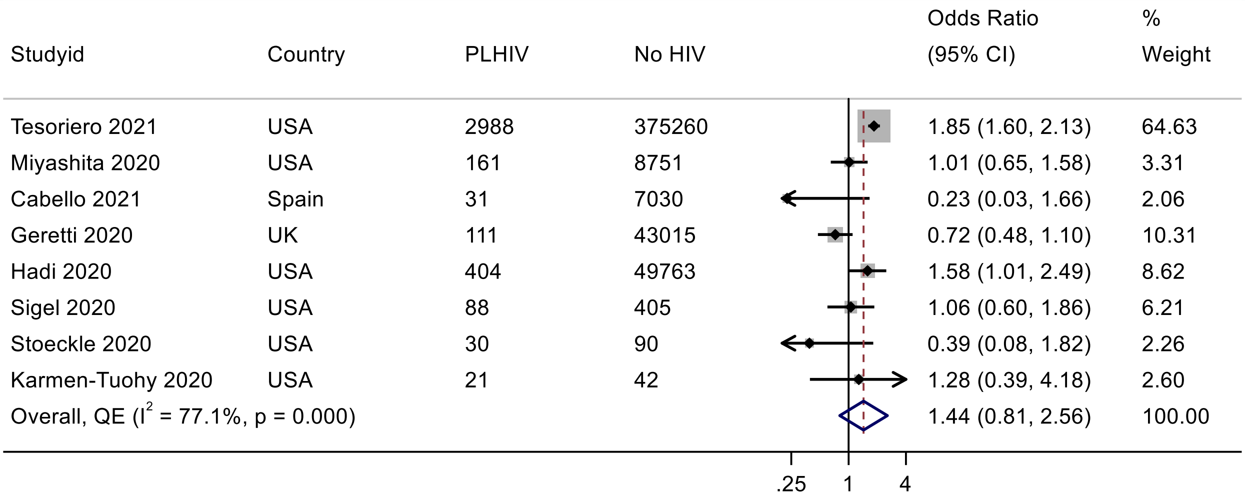

| Studyid | Country | PLHIV | No HIV | Odds Ratio (95% CI) | % Weight |
|---|---|---|---|---|---|
| Tesoriero 2021 | USA | 2988 | 375260 | 1.85 (1.60, 2.13) | 64.63 |
| Miyashita 2020 | USA | 161 | 8751 | 1.01 (0.65, 1.58) | 3.31 |
| Cabello 2021 | Spain | 31 | 7030 | 0.23 (0.03, 1.66) | 2.06 |
| Geretti 2020 | UK | 111 | 43015 | 0.72 (0.48, 1.10) | 10.31 |
| Hadi 2020 | USA | 404 | 49763 | 1.58 (1.01, 2.49) | 8.62 |
| Sigel 2020 | USA | 88 | 405 | 1.06 (0.60, 1.86) | 6.21 |
| Stoeckle 2020 | USA | 30 | 90 | 0.39 (0.08, 1.82) | 2.26 |
| Karmen-Tuohy 2020 | USA | 21 | 42 | 1.28 (0.39, 4.18) | 2.60 |
| Overall, QE ($I^2$ = 77.1%, p = 0.000) | | | | 1.44 (0.81, 2.56) | 100.00 |



NOTE: Weights are from Doi's Quality Effects model

#### Supplementary Fig 4- Hospitalization Doi plot

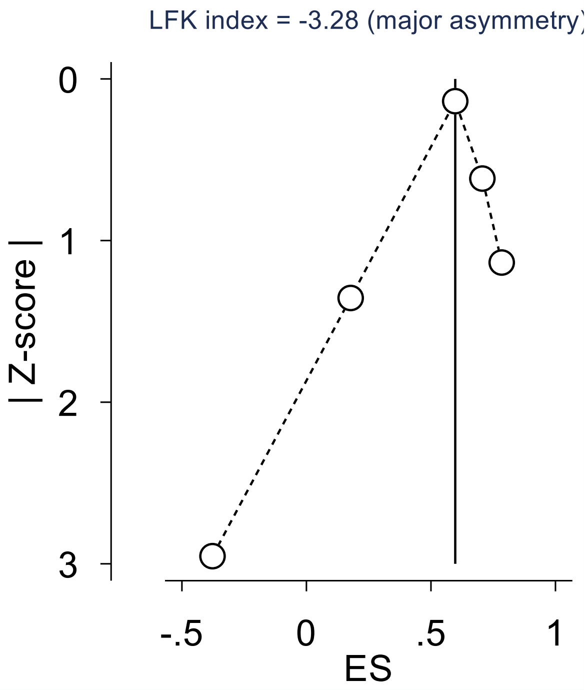

#### Supplementary Fig 5- Odds ratios for hospitalization for COVID-19 by HIV status, without the South Africa studies

| Studyid | Country | PLHIV | No HIV | Odds Ratio (95% CI) | % Weight |
|---|---|---|---|---|---|
| Tesoriero 2021 | USA | 2988 | 375260 | 2.19 (2.02, 2.37) | 40.11 |
| Cabello 2021 | Spain | 31 | 7030 | 0.69 (0.34, 1.39) | 10.82 |
| Hadi 2020 | USA | 404 | 49763 | 2.03 (1.58, 2.60) | 15.07 |
| Kohler 2020 | USA &Spain | 6050 | 752012 | 1.82 (1.72, 1.92) | 34.00 |
| Overall, QE ($I^2$ = 87.0%, p = 0.000) | | | | 1.79 (1.49, 2.15) | 100.00 |

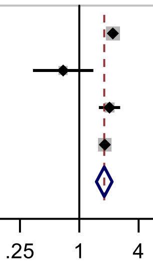


NOTE: Weights are from Doi's Quality Effects model

#### Supplementary Fig 6- Odds ratio for hospitalization by age group

| Age groups and Authors | Country of Study | PLHIV | No HIV | Odds Ratio (95% CI) |
|---|---|---|---|---|
| <40 years | | | | |
| Tesoriero 2021 | USA | 492 | 121871 | 2.25 (1.72, 2.94) |
| 40-60 years | | | | |
| Tesoriero 2021 | USA | 1400 | 133095 | 2.51 (2.22, 2.83) |
| <60 years | | | | |
| Tesoriero 2021 | USA | 1892 | 254966 | 2.82 (2.53, 3.15) |
| ≥60 years | | | | |
| Tesoriero 2021 | USA | 1096 | 119291 | 1.65 (1.46, 1.86) |

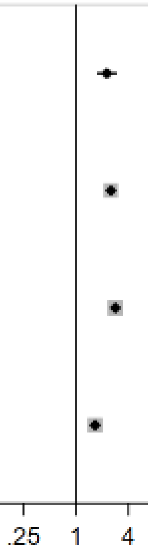

#### Supplementary Fig 7- Intensive care unit Doi plot

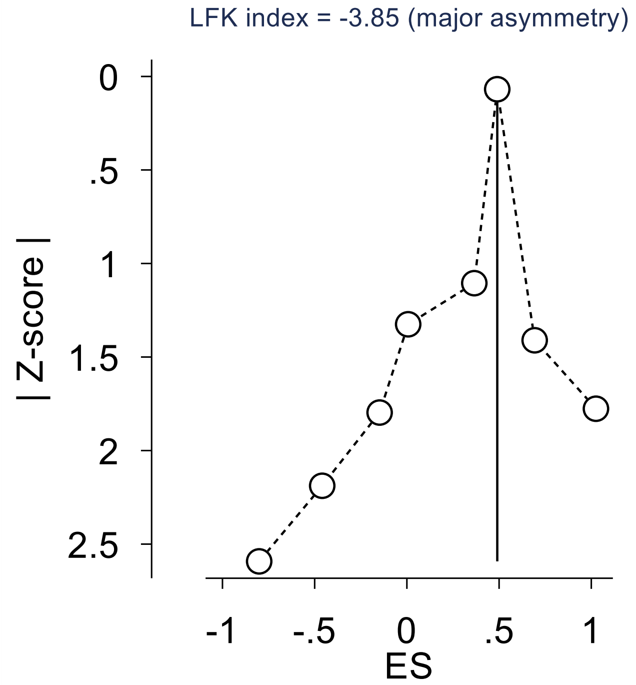


#### Supplementary Fig 8- Odds ratio for intensive care services by age group

| Age groups and Authors | Country of Study | PLHIV | No HIV | Odds Ratio (95% CI) |
|---|---|---|---|---|
| ≤50 years | | | | |
| Miyashita 2020 | USA | 38 | 2981 | 1.73 (0.76, 3.96) |
| >50 years | | | | |
| Miyashita 2020 | USA | 123 | 5770 | 0.80 (0.53, 1.22) |
| 51-65 years | | | | |
| Miyashita 2020 | USA | 82 | 2450 | 1.11 (0.68, 1.81) |
| ≥66 years | | | | |
| Miyashita 2020 | USA | 41 | 3320 | 0.42 (0.18, 1.00) |
| <66 years | | | | |
| Miyashita 2020 | USA | 243 | 11201 | 1.23 (0.94, 1.60) |

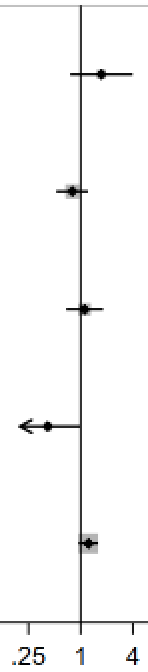

### Supplementary Fig 9 - Comparison of comorbid conditions by HIV status

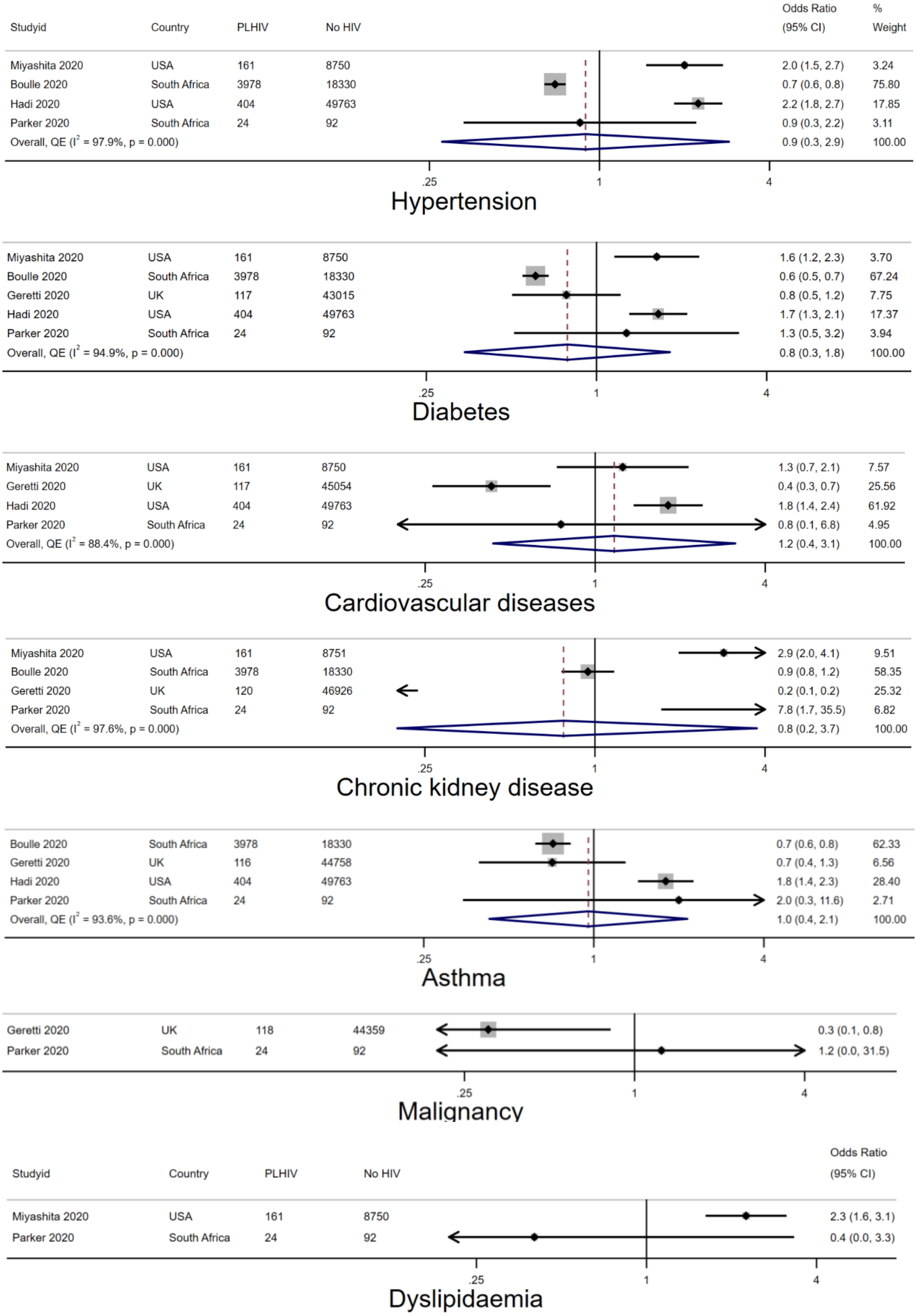

#### Sensitivity analysis – Random effects models

#### Overall mortality

| Studyid | Country | PLHIV | No HIV | Odds Ratio (95% CI) | % Weight |
|---|---|---|---|---|---|
| Tesoriero 2021 | USA | 2988 | 375260 | 1.85 (1.60, 2.13) | 17.07 |
| Miyashita 2020 | USA | 161 | 8751 | 1.01 (0.65, 1.58) | 12.95 |
| Boulle 2020 | South Africa | 3978 | 18330 | 1.04 (0.85, 1.28) | 16.43 |
| Cabello 2021 | Spain | 31 | 7030 | 0.23 (0.03, 1.66) | 2.12 |
| Geretti 2020 | UK | 111 | 43015 | 0.72 (0.48, 1.10) | 13.36 |
| Hadi 2020 | USA | 404 | 49763 | 1.58 (1.01, 2.49) | 12.84 |
| Sigel 2020 | USA | 88 | 405 | 1.06 (0.60, 1.86) | 11.09 |
| Stoeckle 2020 | USA | 30 | 90 | 0.39 (0.08, 1.82) | 3.27 |
| Karmen-Tuohy 2020 | USA | 21 | 42 | 1.28 (0.39, 4.18) | 4.93 |
| Parker 2020 | South Africa | 24 | 92 | 0.89 (0.32, 2.51) | 5.96 |
| Overall, DL ($I^2$ = 78.6%, p = 0.000) | | | | 1.08 (0.79, 1.47) | 100.00 |

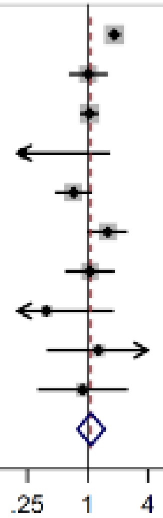


NOTE: Weights are from random-effects model

#### Mortality by age group

| Age categories and Authors | Country of Study | PLHIV | No HIV | Odds Ratio (95% CI) | % Weight |
|---|---|---|---|---|---|
| <60 years | | | | | |
| Boulle 2020 | South Africa | 3826 | 15827 | 1.91 (1.47, 2.48) | 17.68 |
| Tesoriero 2021 | USA | 1892 | 254966 | 4.47 (3.51, 5.68) | 17.79 |
| Miyashita 2020 | USA | 243 | 11201 | 1.09 (0.76, 1.57) | 16.95 |
| Subgroup, DL ($I^2$ = 95.7%, p = 0.000) | | | | 2.12 (0.97, 4.65) | 52.43 |
| ≥60 years | | | | | |
| Boulle 2020 | South Africa | 152 | 2503 | 1.65 (1.09, 2.51) | 16.50 |
| Tesoriero 2021 | USA | 1096 | 119291 | 1.23 (1.03, 1.47) | 18.11 |
| Miyashita 2020 | USA | 41 | 3320 | 0.63 (0.29, 1.37) | 12.97 |
| Subgroup, DL ($I^2$ = 58.2%, p = 0.092) | | | | 1.22 (0.85, 1.75) | 47.57 |
| Overall, DL ($I^2$ = 94.2%, p = 0.000) | | | | 1.58 (0.95, 2.63) | 100.00 |
| Heterogeneity between groups: p = 0.208 | | | | | |

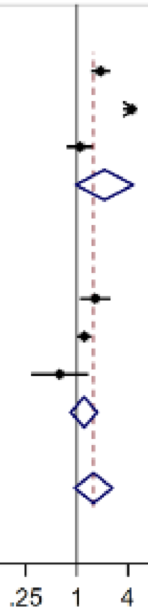


NOTE: Weights and between-subgroup heterogeneity test are from random-effects model

#### Hospitalization

| Studyid | Country | PLHIV | No HIV | Odds Ratio (95% CI) | % Weight |
|---|---|---|---|---|---|
| Tesoriero 2021 | USA | 2988 | 375260 | 2.19 (2.02, 2.37) | 23.99 |
| Boulle 2020 | South Africa | 3978 | 18330 | 1.19 (1.08, 1.32) | 23.70 |
| Cabello 2021 | Spain | 31 | 7030 | 0.69 (0.34, 1.39) | 8.26 |
| Hadi 2020 | USA | 404 | 49763 | 2.03 (1.58, 2.60) | 19.74 |
| Kohler 2020 | USA &Spain | 6050 | 752012 | 1.82 (1.72, 1.92) | 24.30 |
| Overall, DL ($I^2$ = 96.0%, p = 0.000) | | | | 1.62 (1.26, 2.08) | 100.00 |

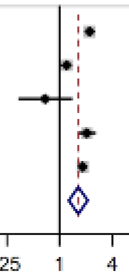


NOTE: Weights are from random-effects model

#### Intensive care services

| Studyid | Country | PLHIV | No HIV | Odds Ratio (95% CI) | % Weight |
|---|---|---|---|---|---|
| Miyashita 2020 | USA | 161 | 8751 | 1.01 (0.69, 1.46) | 16.77 |
| Cabello 2021 | Spain | 31 | 7030 | 1.44 (0.34, 6.07) | 4.32 |
| Geretti 2020 | UK | 122 | 46497 | 2.79 (1.90, 4.08) | 16.64 |
| Kohler 2020 | USA &Spain | 5762 | 630159 | 1.63 (1.45, 1.84) | 20.66 |
| Farina 2020 | South Africa | 55 | 69 | 1.16 (0.53, 2.57) | 9.70 |
| Sigel 2020 | USA | 88 | 405 | 0.86 (0.49, 1.52) | 13.18 |
| Stoeckle 2020 | USA | 30 | 90 | 0.63 (0.22, 1.85) | 6.66 |
| Karmen-Tuohy 2020 | USA | 21 | 42 | 2.00 (0.57, 6.96) | 5.38 |
| Parker 2020 | South Africa | 24 | 92 | 0.45 (0.15, 1.31) | 6.68 |
| Overall, DL ($I^2$ = 70.9%, p = 0.001) | | | | 1.27 (0.91, 1.77) | 100.00 |

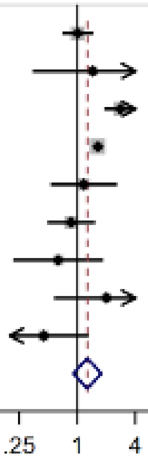


NOTE: Weights are from random-effects model
